# Design and outcome-model estimability in doubly robust survival analysis: a breast cancer reanalysis

**DOI:** 10.64898/2026.09.29.26364358

**Authors:** Wael Fahmy, Elena Krikun, Abedalrhman Alkhateeb

## Abstract

Doubly robust survival estimators require nuisance models that the available information can support. We examine design and estimator dependence in TCGA breast cancer data. Treatment indicators disagree for 337 of 1076 patients; 67.4% have unobserved five-year binary outcomes; and receptor imputation expands the triple-negative subgroup from 156 directly measured patients to 226. We compare chemotherapy initiation by day 90 with non-initiation, using adjusted restricted mean survival time (RMST) over the next 1095 days. Weighting-only estimates are 55 days (95% bootstrap interval: −24, 133) in triple-negative disease (16 deaths) and 38 days (12, 72) in the whole cohort (72 deaths). A Cox outcome-model benchmark yields 108 and 35 days, respectively, demonstrating material model dependence in the smaller population. A parameter-to-information ratio summarizes the burden on complete-outcome regressions within cross-fitting. Its simulation-calibrated warning level is 0.31 (0.28–0.35) for the original regime; additional simulations examine covariate-dependent censoring, delayed benefit and nonlinear misspecification. The ratio does not certify estimator validity or replace weight diagnostics. The contribution is a reproducible workflow that separates design, nuisance-model support and estimator stability before interpreting observational survival contrasts.

## 1. Introduction

Public cancer cohorts contain detailed molecular measurements, yet their clinical records do not always support equally detailed treatment questions. The Cancer Genome Atlas breast cancer cohort (TCGA-BRCA), for example, was assembled to characterize tumors rather than to compare treatment strategies (Cancer Genome Atlas Network, 2012). Before asking whether chemotherapy is associated with longer survival, an analyst must establish what the treatment field records, when treatment began, and which patients have sufficient follow-up to contribute to the chosen outcome. These decisions determine the comparison that a statistical model can estimate.

Modern causal methods do not remove this dependence on study design. Double/debiased machine learning separates estimation of a target parameter from estimation of nuisance functions, with cross-fitting helping to control the effects of fitting flexible models (Chernozhukov et al., 2018). Doubly robust survival estimators combine treatment and outcome models to reduce reliance on either model alone under specified assumptions (Bai, Tsiatis, and O’Brien, 2013; Zhang and Schaubel, 2012). These properties do not repair an ambiguous exposure or an outcome assigned to a patient whose status is unobserved. Finite-sample performance also depends on whether the data can support the nuisance models being fitted.

These problems are concrete in TCGA-BRCA. Two treatment indicators disagree for 337 of 1076 patients; a historical five-year mortality indicator assigns survival to 725 patients censored before that horizon; and imputed receptor status expands the triple-negative breast cancer (TNBC) subgroup from 156 directly measured patients to 226. We use a day-90 landmark to make treatment classification and the start of follow-up explicit (Dafni, 2011; Hernán et al., 2016). Restricted mean survival time (RMST), the area under the survival curve over a specified period, expresses the comparison in days without assigning a binary outcome to patients with incomplete follow-up (Conner et al., 2019).

The study builds on earlier multimodal feature-selection work (Krikun and Alkhateeb, 2026b). A covariate representation supplied from an earlier analysis is used here as one of three adjustment specifications, not as a newly discovered molecular signature. Prognostic feature selection and confounder selection answer different questions (VanderWeele, 2019); the published feature-selection framework is not re-evaluated in this study.

We examine how treatment, subgroup and time-origin choices change the analysis, compare four survival estimators on a common RMST scale, and assess whether limited outcome-model information helps explain estimator sensitivity. Unadjusted Kaplan–Meier, weighted Kaplan–Meier, cross-fitted augmented inverse probability weighting (AIPW), and a standardized Cox outcome model provide complementary comparisons. Our diagnostic contribution is a parameter-to-information ratio for the complete-outcome regression inside AIPW. Unlike prediction-model sample-size guidance (Peduzzi et al., 1996; Riley et al., 2019; van Smeden et al., 2016), its calibration concerns adjustment-set sensitivity relative to sampling variability. It complements study-specific simulation, including approaches such as REFINE2 (Meng and Huang, 2026), rather than providing a universal sample-size rule. Independent stress scenarios examine the limits of the original calibration. The objective is estimation and assessment of a population-level survival contrast, not individual prediction, classification or clustering.

## 2. Materials and Methods

### 2.1 Study workflow and data sources

Figure 1 summarizes the workflow from the clinical-record audit to the interpretation of survival contrasts. We use archived NCI Genomic Data Commons clinical and follow-up exports (Grossman et al., 2016) and an earlier derived UCSC Xena overall-survival table (Goldman et al., 2020). The supplied survival table is reconciled with clinical records; the original Xena export and original high-dimensional omics matrices are not included. The earlier multimodal adjustment set is supplied as a derived input, not reselected in this study. Web Appendices B and I describe the exclusion ledger and data provenance.

**Figure 1.**
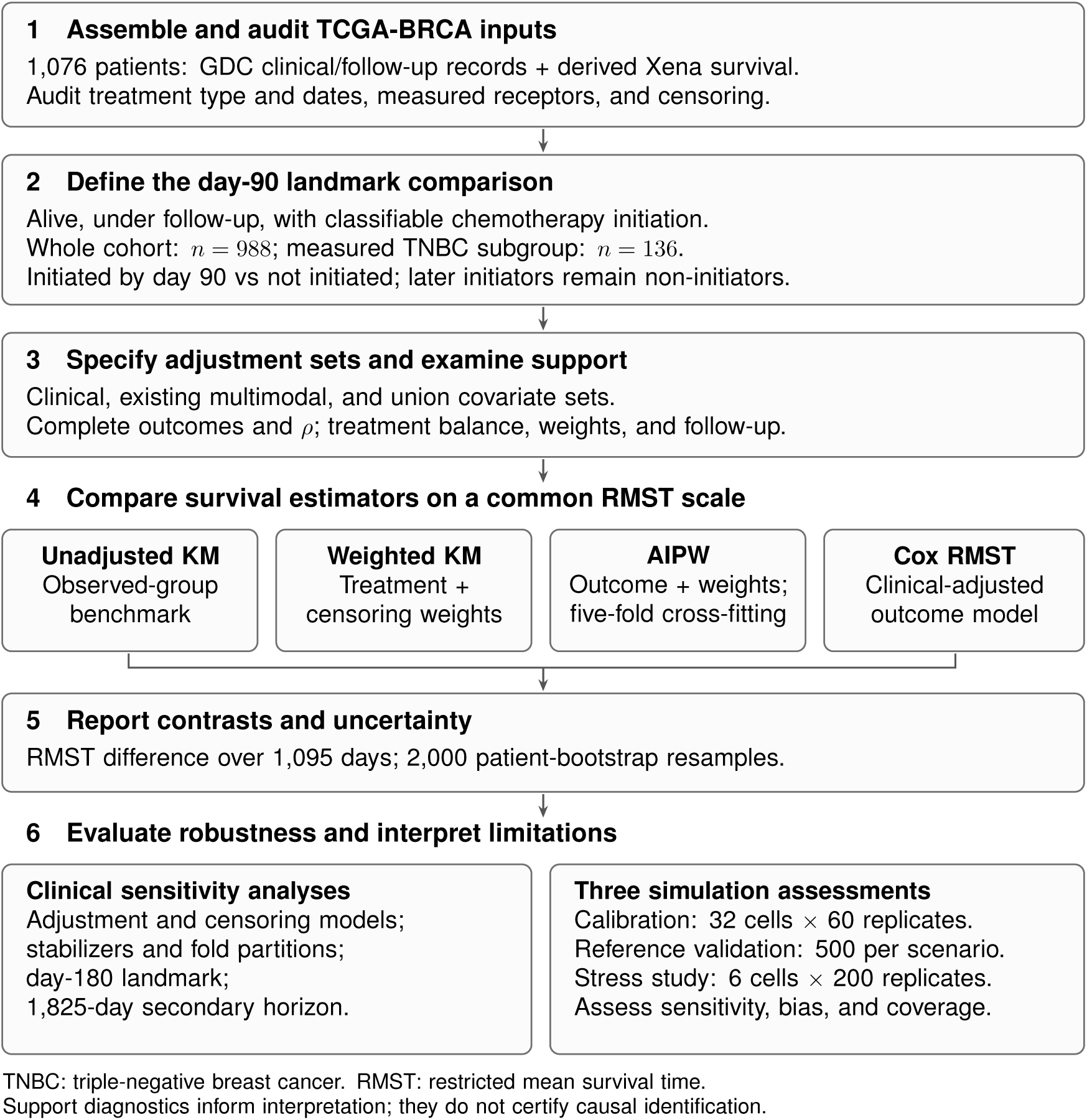
Study workflow. The information ratio, balance and follow-up support inform interpretation; they do not automatically select an estimator or establish causal identification. Initiation status is fixed at the landmark; the workflow does not imply prospective registration. *Alt text:* The flow links 1,076 input records to 988 eligible patients and a 136-patient TNBC subgroup, four survival estimators, RMST contrasts, and clinical and simulation assessments.

Chemotherapy initiation is examined in TNBC and in the whole eligible cohort, without designating either population primary. TNBC provides a more clinically specific comparison but has little event information. The whole-cohort analysis offers more information while pooling receptor subtypes with different treatment indications. Hormone therapy is a different treatment question and is not estimated here; its recipients are not thereby excluded from the whole-cohort chemotherapy analysis.

### 2.2 Treatment, receptor status and landmark eligibility

Chemotherapy and its first recorded start are derived from the structured treatments. treatment_type and treatments.days_to_treatment_start fields. The earlier indicator used the serialized treatments.treatment_or_therapy field. The audit establishes discordance between classifications, not an externally validated error rate.

Estrogen, progesterone and HER2 receptor records retain positive, negative, equivocal, conflicting and untested states. TNBC requires all three receptors to be directly recorded as negative; cohort-defining receptors are never imputed. Indeterminate receptors exclude patients from the corresponding subgroup, but not automatically from the whole-cohort analysis.

Patients who die or are censored before day 90 are excluded, as are chemotherapy recipients without a usable initiation date. Exposure is fixed at day 90 and follow-up is reset to that date. Later initiators remain in the non-initiator group. A secondary analysis repeats this construction at day 180. Table 1 reports the primary analysis populations and information available for estimation.

**Table 1.** Information and support at the day-90 landmark, clinical adjustment set and 1095-day horizon. Complete outcomes include observed deaths and follow-up reaching the horizon; they are not event counts. Effective degrees of freedom refer to the smaller complete-outcome arm across five training folds. SMD denotes absolute standardized mean difference under primary weighted-KM treatment weights.

| Quantity | TNBC | Whole cohort |
| --- | --- | --- |
| Patients | 136 | 988 |
| Chemotherapy initiators / non-initiators | 90 / 46 | 400 / 588 |
| Deaths within the primary horizon | 16 | 72 |
| Complete outcomes, initiators / non-initiators | 44 / 20 | 189 / 264 |
| Retained covariate columns | 24 | 27 |
| Nominal outcome parameters, with intercept | 25 | 28 |
| Clinical information ratio $\rho$ | 1.56 | 0.19 |
| Smaller-arm training-fit effective degrees of freedom | 6.8–7.5 | 17.1–18.4 |
| Mean SMD before / after treatment weighting | 0.173 / 0.113 | 0.173 / 0.051 |
| Maximum SMD after treatment weighting | 0.379 | 0.196 |

### 2.3 Target contrast and assumptions

Let *L* = 1 denote eligibility for the analysis at the day-90 landmark, including being alive, under follow-up and having classifiable treatment status; TNBC additionally requires measured receptors. Let *A* indicate chemotherapy initiation by day 90 and *X* denote the adjustment covariates. Survival and censoring times from the landmark are *T* and *C*; we

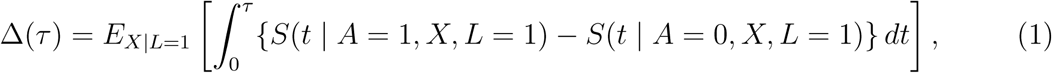

with *τ* = 1095 days primary and 1825 days secondary. A positive contrast indicates longer restricted mean survival among initiators after standardization to the eligible population. It compares initiation status by the landmark, not chemotherapy with a sustained no-chemotherapy strategy. Identification requires treatment positivity, positive censoring survival through the horizon, and *C ⊥ T | A, X, L* = 1. Causal interpretation additionally requires consistency for a sufficiently specified treatment history and conditional exchange-ability (Hernán and Robins, 2020).

Landmarking avoids crediting follow-up before classification to the treated group, but does not randomize treatment. Treated patients are prevalent users (Ray, 2003); early treatment may affect reaching the landmark. Conditioning on eligibility can therefore induce selection. Neither the number excluded nor a second landmark bounds this bias. The analysis does not emulate a trial assigning treatment at diagnosis or among untreated patients at day 90.

### 2.4 Adjustment sets and fixed preprocessing

The primary clinical adjustment set represents age, stage, T, N and M category, calendar era and contributing site. Site can proxy case mix, treatment practice, care and follow-up; association with treatment and outcome alone does not establish confounding (VanderWeele, 2019). Grade is entirely missing and a tumor-size field is unavailable; T category is a coarse proxy. The earlier multimodal set and its union with the clinical set are sensitivity specifications. After constant columns are removed, the clinical matrices contain 24 covariate columns in TNBC and 27 in the whole cohort; adding an intercept gives 25 and 28 nominal outcome-model parameters. Prognostic feature selection does not establish a valid confounder set.

Clinical continuous and ordinal missing values use median replacement with missingness indicators; stage and TNM categories have fixed ordinal coding, and small sites are pooled. The supplied earlier feature matrix uses its archived preprocessing, with remaining nonfinite values replaced as documented in Web Appendix A. Clinical encoding, imputation and the supplied feature set are held fixed for the reported resampling analysis. For AIPW, propensity/outcome scaling is computed on the analysis matrix, while fitted regression coefficients and censoring-model training use the training folds. This is not a fully nested feature-learning or preprocessing pipeline. The clinical-first and weighting-only-first orderings were chosen after estimates were available; they were not prospectively registered. Both estimators and all adjustment sets are retained.

### 2.5 Survival estimators and comparator design

The primary estimator combines stabilized treatment weights with time-varying inverse censoring weights in a Kaplan–Meier product limit (Conner et al., 2019; Robins and Finkelstein, 2000). Write *e*(*X*) = Pr(*A* = 1 *| X, L* = 1), *G*(*t | A, X*) = Pr(*C > t | A, X, L* = 1) and *w_i_* = Pr(*A* = *A_i_ | L* = 1)*/* Pr(*A* = *A_i_ | X_i_, L* = 1). At the observed event times *t_j_* in arm *a*,

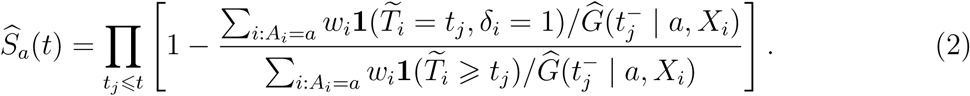

Integrating the two curves gives an RMST contrast. Propensity is ridge logistic; censoring uses a ridge Cox model stratified by treatment with shared covariate coefficients. Both are fitted on the full analysis sample for this estimator. An arm-only censoring curve cancels within each risk-set ratio, whereas covariate-conditional censoring weights do not. Weighting-only estimation is not doubly robust.

The sensitivity estimator is a five-fold cross-fitted AIPW estimator of a complete restricted outcome: death observed by *τ*, or follow-up reaching *τ*. Outcome regressions are IPCW-weighted ridge least squares fitted within each treatment arm. Before truncation, its equation is doubly robust in propensity and outcome regression conditional on a correct censoring model. It omits the additional censoring augmentation used in more general survival estimators (Bai et al., 2013; Zhang and Schaubel, 2012); Web Appendix A supplies the equation and expectation identity. Cross-fitting alone does not establish nuisance-rate conditions or eliminate regularization bias.

For clarity, write *Y_i_*= min(*T̃_i_, τ*), let *D_i_* indicate a complete restricted outcome, and let *m̂_a,−k_*_(_*_i_*_)_(*X_i_*) be the outcome regression trained without patient *i*’s fold. Before stabilization, the estimator averages

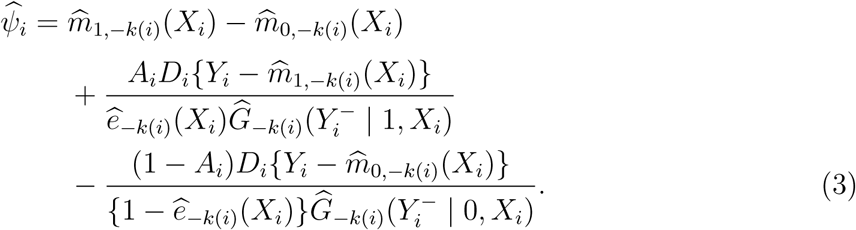

The augmentation adds a weighted residual correction to the difference between the two outcome-model predictions. When complete outcomes are scarce in one treatment arm, these predictions may depend strongly on regularization and the adjustment set. This motivates the information diagnostic below without implying that weighting-only estimation is preferable. Two benchmarks address different aspects of model dependence. Unadjusted Kaplan–Meier RMST (Kaplan and Meier, 1958) describes the observed treatment groups without confounder control. The adjusted outcome benchmark fits *h*(*t | A, X*) = *h*_0_(*t*) exp(*θA*+*β^⊤^X*) (Cox, 1972), with an unpenalized treatment coefficient and ridge-penalized standardized covariates. Its predicted survival under each treatment is standardized over the same analysis population and integrated to the same *τ*. Thus its comparison with the adjusted estimators is in RMST days, not between a hazard ratio and a mean difference. This death-hazard model is distinct from the Cox censoring model used in weighting.

Propensity limits are 0.01/0.99 for weighted KM and 0.02/0.98 for AIPW; censoring survival is floored at 0.05. AIPW caps positive combined inverse weights at the 99th percentile and clips outcome predictions and the final contrast to feasible ranges. These devices can introduce bias. We vary each separately and jointly. The ridge rule, clipping conventions, weighting diagnostics and exploratory proportional-hazards screens are specified in Web Appendices A, C, D and G. Covariate balance is assessed using standardized mean differences under each estimator’s treatment weights (Austin and Stuart, 2015). Schoenfeld residual screens (Schoenfeld, 1982) use within-model Holm adjustment (Holm, 1979); they are exploratory, not a proof of model adequacy.

### 2.6 Information diagnostic and simulation assessment

For *p* outcome-model parameters, including the intercept after constant columns are removed, *K* folds and *c*_min_ complete restricted outcomes in the smaller treatment arm, define

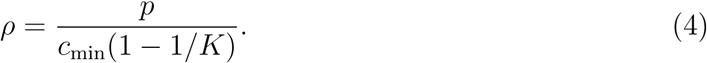

The denominator is an expected training-fold count; actual partitions differ. The diagnostic precedes effect-model fitting but is not outcome-blind because completeness depends on follow-up and event status. It applies to this complete-outcome regression, not automatically to survival learners that use censored observations differently. We report effective degrees of freedom from the actual Cox-weighted outcome fits beside nominal dimension.

The original calibration has 32 cells crossing sample sizes 150, 300, 600 and 1000, covariate dimensions 10, 25, 50 and 75, and two event-rate settings, with 60 replicates per cell. Covariates are Gaussian, treatment is logistic, survival exponential and censoring covariate-independent. The first three covariates contain the treatment-assignment predictors, and all three nested adjustment sets retain them. The calibration target is the mean within-replicate range across adjustment sets divided by the full-set estimator’s sampling standard deviation. A log-linear fit against *ρ* defines the crossing at 0.50, a chosen warning convention. Web Appendix F gives the exact calibration response, its aggregation across replicates, cell-bootstrap intervals, alternative crossing levels and penalty sensitivities. This is not a calibration to acceptable bias or coverage.

We add six independent stress cells: sample sizes 136 and 988 with 24 and 27 covariates, respectively, crossed with covariate-dependent censoring under proportional hazards, delayed treatment benefit, and heavy-tailed covariates with nonlinear propensity, survival and censoring mechanisms. Each cell uses 200 replicates and 199 inner patient bootstraps. Baseline hazards are calibrated to expected event counts near 16 or 72. Analytic conditional RMST provides the replicate’s covariate-standardized truth. We compare all four estimators using bias, root mean squared error (RMSE), interval coverage, standard-error calibration, failures and adjustment-set sensitivity. Monte Carlo uncertainty is reported, including Wilson intervals for coverage. These scenarios test behavior outside the original grid; they do not supply external clinical validation. Web Appendix G gives all mechanisms and results.

### 2.7 Uncertainty and sensitivity analyses

Clinical contrast intervals are patient-level percentile bootstrap intervals from 2000 replicates, refitting the estimators on each resampled fixed covariate matrix. Percentile intervals are not constrained to be symmetric about the point estimate. New KM/Cox benchmarks use the same bootstrap draws as each other; existing estimators retain their documented seeds. No synthetic oversampling is used. The original operating-point simulation uses 500 replicates per scenario with 500 inner bootstraps, and is retained separately from the new stress study. Additional checks vary adjustment sets, censoring models, stabilizers, landmark and horizon, and repeat AIPW over 50 partitions and the core analysis over six seeds. The correction ladder changes one design component at a time on a common RMST scale; its rungs need not have an unchanged target. Exploratory heterogeneity is retained descriptively in Web Appendix H; its earlier fixed-score intervals do not support confirmatory inference.

## 3. Results

### 3.1 Cohort construction and observed follow-up

Of 1076 patients, 57 do not reach the landmark, 29 have chemotherapy without a usable start date, and 2 have implausible prediagnosis starts, leaving 988. Of these, 400 initiated chemotherapy by day 90 and 145 initiate later. TNBC contains 136 patients, with 90 initiators and 46 non-initiators. Deaths within the primary horizon number 16 and 72 in TNBC and the whole cohort. Only 6 HER2-positive patients initiated targeted therapy by the landmark; no targeted-therapy contrast is estimated.

Median reverse-Kaplan–Meier follow-up in the original cohort is 974 days. At five years, 201 patients are observed alive without a subsequent recorded death, 100 died by that horizon, 50 die later and therefore survived the horizon, and 725 are censored earlier. Only the last group has unknown five-year status. Coding them as survivors is unjustified even under independent censoring. Web Appendix B and Web Figures 1–2 present the cohort ledger and follow-up audit.

**Figure 2.**
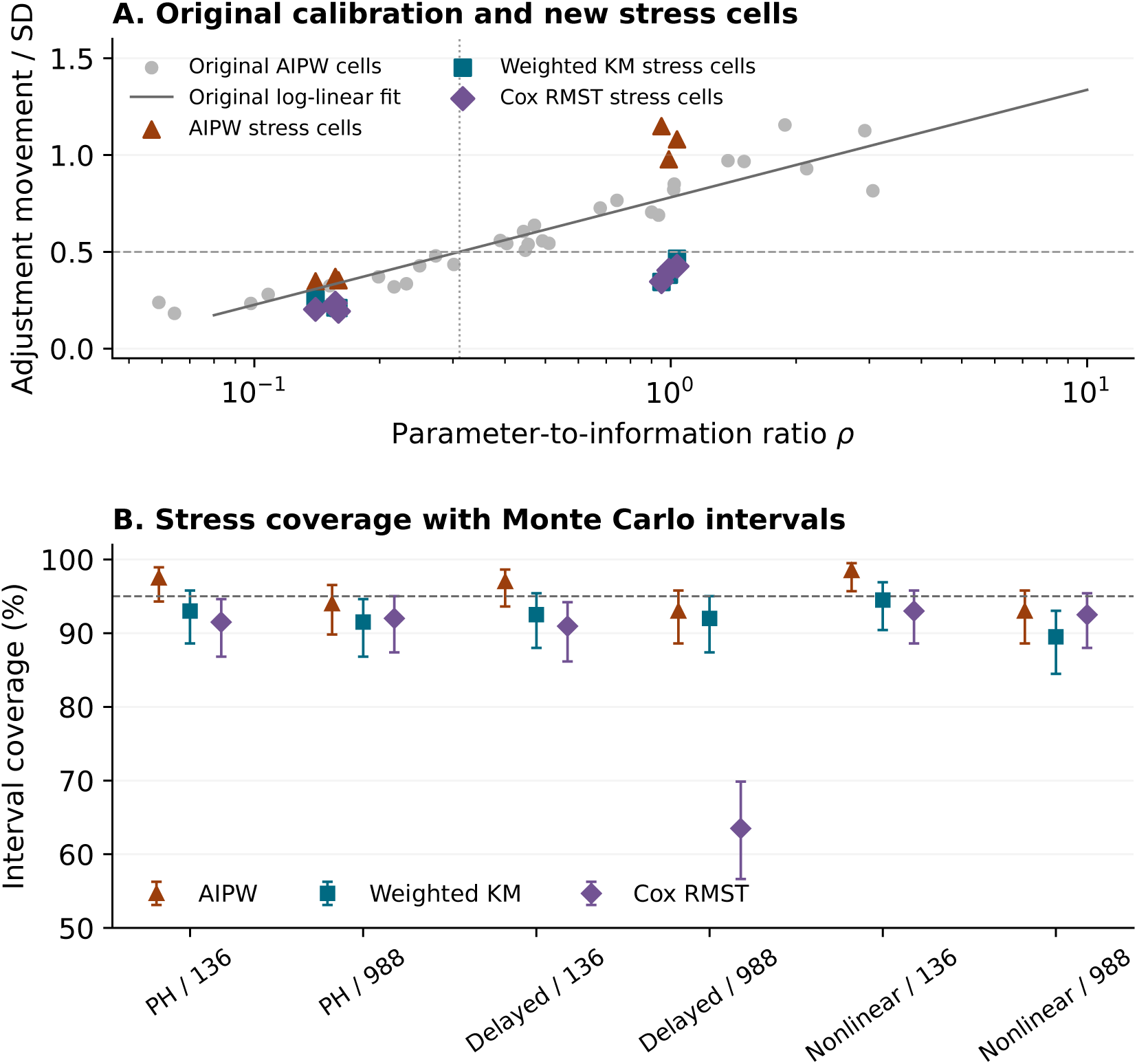
Original calibration and independent stress assessment. The original log-linear AIPW fit and its 0.50 crossing are shown with the original grid cells and new scenario means. The lower panel gives coverage of nominal 95% bootstrap intervals in the stress study; bars are 95% Wilson intervals for Monte Carlo uncertainty. These assess the simulated settings, not clinical coverage. Weighted KM and AIPW denote the adjusted implementations; Cox uses standardized outcome-model survival. *Alt text:* Calibration points appear above simulated coverage intervals. In the larger delayed-benefit cell, Cox coverage is 63.5 percent, compared with 92 percent for weighted KM and 93 percent for AIPW.

### 3.2 Same-horizon estimator comparison

Table 2 gives all four estimators under the clinical specification. In TNBC, weighting-only estimation gives 55.0 days (95% interval: −24.4, 132.5), and AIPW gives 19.5 (−118.5, 179.7). The standardized Cox estimate is 108.3 (30.5, 167.6), compared with unadjusted KM 54.6 (−29.1, 144.7). The Cox interval excludes zero while the weighted-KM and AIPW intervals span zero. This disagreement shows model dependence on 16 deaths, not independent confirmation of benefit.

**Table 2.**
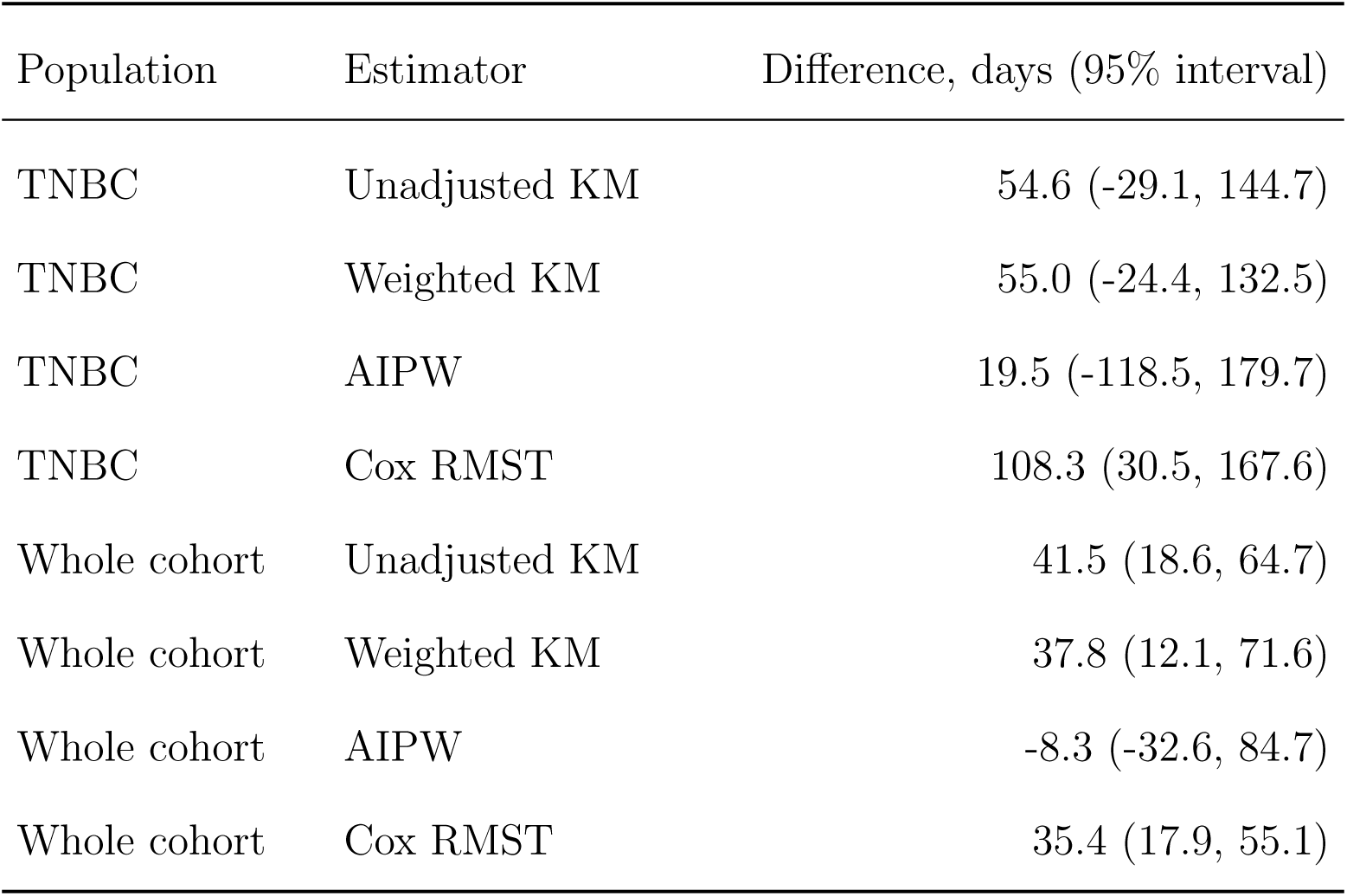
RMST contrasts over 1095 days after the day-90 landmark. Intervals are 95% patient-bootstrap percentile intervals. The unadjusted benchmark compares observed groups; the adjusted methods target the standardized contrast under their respective assumptions. Cox denotes an outcome model.

In the whole cohort, weighted KM gives 37.8 (12.1, 71.6), Cox 35.4 (17.9, 55.1), and AIPW −8.3 (−32.6, 84.7). The Cox and weighted-KM point estimates are close, while AIPW changes sign. The unadjusted estimate is 41.5 (18.6, 64.7). Each adjusted method relies on different modeling assumptions; agreement does not prove absence of confounding.

Across adjustment sets, TNBC weighted-KM estimates range from 28.6 to 55.0 days, and AIPW from −5.4 to 42.0. Changing covariate content changes nuisance fits and confounding control, so the difference cannot be attributed to parameter count alone. At the secondary TNBC horizon, weighted KM gives 256 (32, 516) days and AIPW 384 (107, 670) days; both intervals exclude zero, but tail support is limited. The day-180 landmark gives whole-cohort AIPW 26.5 rather than −8.3 days; both intervals include zero. Web Tables 2 and 6 retain the complete results; Web Figure 5 displays the original two-estimator comparison across adjustment sets.

### 3.3 Information, balance and censoring support

TNBC has 44/20 complete outcomes in initiators/non-initiators, 25 nominal outcome-model parameters and *ρ* = 1.56. The whole cohort has 189/264 complete outcomes in initiators/non-initiators, 28 parameters and *ρ* = 0.19. In the smaller complete-outcome arm, actual training-fit effective degrees of freedom range 6.8–7.5 in TNBC and 17.1–18.4 in the whole cohort. These are computed from the fitted cross-fold Cox-weighted regressions, not from an arm-only reference fit.

Under the primary KM treatment weights, mean absolute standardized differences change from 0.173 to 0.113 in TNBC and from 0.173 to 0.051 in the whole cohort; the corresponding maximum post-weighting differences are 0.379 and 0.196. AIPW’s capped complete-outcome residual-weight effective sample sizes are 31/7 and 77/86 for treated/control patients. These are estimator-specific weight summaries, not a universal sample size or a direct variance formula for weighted KM. Full balance and support diagnostics accompany the code.

Across 15 named sites with at least 20 patients, median follow-up ranges from 293 to 5086 days; smaller sites are pooled into an OTHER category for modeling. The censoring floor binds on 0/4 complete cases in the two weighted-KM fits and 2/7 in cross-fitted AIPW. Exploratory Schoenfeld screens of Efron-tie reference refits identify one covariate–process combination after within-model Holm adjustment: diagnosis year (year dx) in the whole cohort censoring model (adjusted *p* = 0.0023). Penalization and few deaths limit these screens, and nonsignificance cannot establish proportional hazards or independent censoring. Web Table 11 reports every tested term.

The revised five-device stabilizer analysis moves AIPW by up to 80.7 days; weighted-KM shifts reach 10.2 days (Web Table 5). Across 50 partitions, TNBC AIPW ranges from −134 to 81 days and whole-cohort AIPW from −19 to 29 days. Across the six-seed repeat, whole-cohort weighted-KM intervals exclude zero and AIPW intervals span zero; both TNBC methods retain intervals spanning zero. Thus computational reproducibility under one seed does not imply partition stability.

### 3.4 Calibration and independent stress assessment

In the original grid, sensitivity is approximately 0.782 + 0.241 log *ρ* with *R*^2^ = 0.900. The 0.50 crossing is 0.31 (cell-bootstrap interval 0.28–0.35). Changing the crossing to 0.25 or 1.00 yields 0.11 or 2.47; penalty multipliers 0.3 and 3 yield 0.22 or 1.32. Figure 2 juxtaposes the original calibration with the independent stress cells. The maximum original-grid weighted-KM sensitivity ratio is 0.47, and the median AIPW/KM movement ratio is 2.2. These are results for the tested implementations, not a general ranking.

The independent stress study comprised six cells, 1200 simulated samples and 4800 estimator–replicate evaluations. With proportional hazards and covariate-dependent censoring, coverage across the adjusted methods and two sample sizes ranged from 91.5 to 97.5%, with absolute bias up to 14.0 days. Under delayed treatment benefit, coverage ranged from 63.5 to 97.0%, with absolute bias up to 19.2 days. Under nonlinear heavy-tailed misspecification, the corresponding ranges were 89.5–98.5% and up to 5.7 days.

The most pronounced coverage failure occurred for Cox-standardized RMST at *n* = 988 under delayed benefit: 63.5% (95% Monte Carlo interval 56.6–69.9%), compared with 92.0% for weighted KM and 93.0% for AIPW in the same cell. The comparison illustrates a limitation of the constant-hazard-ratio outcome model in that scenario, rather than a general disadvantage of Cox regression. One Cox estimator–replicate evaluation, in the smaller delayed-benefit cell, failed the evaluability criterion. Coverage is conditional on successful evaluations, and failures are reported separately.

Web Table 10 reports all four methods, including the unadjusted benchmark, with bias, RMSE and Monte Carlo uncertainty. The additional sensitivity points in Figure 2 test the warning relationship outside the original grid; they do not recalibrate its crossing. Simulation therefore separates model dependence from clinical identification: a method can perform well against known truth in the tested scenarios without establishing that the clinical comparison is free from selection or confounding.

The original validation simulation gave coverage 92.2–95.8% with absolute bias up to 12.5 days. Its distributions and covariate dimensions approximate aspects of the application; they do not reproduce all site, missingness or treatment-selection processes. Neither simulation study establishes the coverage of the clinical intervals.

### 3.5 Design correction ladder

Table 3 retains every distinct RMST rung: 26.5, 40.1, 42.2, 54.6, 55.0 days. The full progression is 28.6 days, calculated from the unrounded estimates, compared with 35.5 days between weighted KM and AIPW in the final TNBC clinical specification. The cohort contracts from 226 to 136. The original censoring-blind binary risk differences are −7.6, −10.3, −9.1 percentage points; they are an audit of the earlier endpoint and are not differenced against RMST. Successive rungs change treatment classification, eligibility or time zero, so the progression is not an additive decomposition of bias for one fixed estimand. Its magnitude is of similar order to estimator choice.

**Table 3.** TNBC design correction ladder on a common 1095-day RMST scale. Rungs 0–3 are unadjusted and rung 4 uses clinical weighted KM. Event counts use each rung’s time origin. Changing the binary endpoint to RMST is a change of target, not an additional numerical rung. Each interval belongs to its own specification. The day-90 landmark row and the unadjusted KM estimate in Table 2 share the same point estimate (54.6 days), but their interval endpoints differ slightly because they use different documented sets of 2000 bootstrap draws.

| Rung | Specification | *n* | Deaths | RMST (95% interval) |
| --- | --- | --- | --- | --- |
| 0 | Original coding and imputed TNBC | 226 | 25 | 26.5 (-28.6, 84.8) |
| 1 | Structured chemotherapy field | 226 | 25 | 40.1 (-21.0, 105.9) |
| 2 | Measured receptors only | 156 | 15 | 42.2 (-56.5, 164.8) |
| 3 | Day-90 landmark | 136 | 16 | 54.6 (-35.6, 154.5) |
| 4 | Clinical adjustment | 136 | 16 | 55.0 (-24.4, 132.5) |

TNBC: triple-negative breast cancer. RMST: restricted mean survival time. Support diagnostics inform interpretation; they do not certify causal identification.

## 4. Discussion

The reanalysis separates three questions: whether the design defines a coherent comparison, whether the outcome nuisance model has adequate information, and whether treatment and censoring weights have support. The added Cox benchmark demonstrates why a conventional survival model is useful as a comparator but cannot adjudicate the observational answer. Its TNBC interval excludes zero while the other adjusted estimators disagree. In the whole cohort, weighted KM and Cox agree more closely, yet the cross-fitted AIPW estimate is sensitive to weights and partitions.

The ratio *ρ* describes one information burden. A high ratio should prompt simpler specifications, fold-level checks or study-specific simulation; it neither proves invalidity nor selects weighted KM. A low ratio cannot certify overlap, censoring support or stable weights. Its warning level depends on the chosen crossing, penalties and data-generating regime. The independent stress study probes those limits using known RMST truth, but a small collection of scenarios does not establish a universal boundary. Effective complexity, stronger censoring diagnostics and other outcome learners require further calibration.

Clinical interpretation remains limited by prevalent treatment use, treatment-dependent landmark eligibility and substantial scope for confounding by indication. Performance status, comorbidity, preference, grade and size are incompletely measured or absent. Receptor records cannot resolve assay modality or clinically adjudicate conflicts, and treatment records lack external validation. Some sites offer little follow-up at the chosen horizon; truncation trades instability for potential bias. Covariate preprocessing and model order were not prospectively fixed, and the reported bootstrap conditions on the constructed feature matrix. The two populations overlap and the whole cohort pools distinct treatment indications.

This is a treatment-contrast estimation study. It does not develop a disease classifier, clustering procedure or new molecular signature. Bias, coverage, RMSE and support therefore assess its statistical question; classification accuracy or clustering indices would assess a different target. The molecular variables in the inherited adjustment representation were not selected here as a biological signature. GO/KEGG or other functional enrichment of that adjustment set could describe the provenance of included molecular covariates, but it would not validate Δ(*τ*), the treatment contrast, or the identifying assumptions. Independent clinical validation would require dated treatment initiation, appropriately timed covariates, measured receptors and adequate follow-up. Public treatment indicators alone are insufficient for reproducing the landmark design.

The published feature-selection framework (Krikun and Alkhateeb, 2026b) provides context for the earlier adjustment representation, not external validation of this treatment contrast. The related sequencing study (Krikun and Alkhateeb, 2026a) examines hormone-therapy initiation by day 180 in HR-positive, HER2-negative disease and the influence of upstream chemotherapy on a 730-day overlap-population contrast. The separate TCGA– METABRIC study (Krikun and Alkhateeb, 2026c) evaluates prognostic transport and incremental predictive utility. These studies share TCGA breast cancer data and elements of the earlier cohort construction, but address different targets. Web Appendix J describes the scope of the related work and of the archived implementation audit.

## 5. Conclusion

A useful observational survival analysis must make three components visible: the comparison defined by the design, the information available for nuisance-model fitting, and the sensitivity of the estimates to modeling choices. In this reanalysis, design corrections and estimator choice both materially change the RMST contrast. The information ratio identifies one potential source of instability, while weight, partition and simulation diagnostics reveal limitations it cannot detect. These findings support specifying the target and analysis ordering before inspecting contrasts and reporting disagreement rather than selecting the most favorable estimate. They do not establish individual treatment benefit or resolve chemotherapy effectiveness.

## Acknowledgments

The results use data generated by the TCGA Research Network.

## Funding

This work received no specific grant from public, commercial or not-for-profit funding agencies.

## Conflict of interest

The authors declare no conflicts of interest.

## CRediT author statement

W.F.: Conceptualization; Methodology; Software; Formal analysis; Validation; Visualization; Writing–original draft. E.K.: Conceptualization; Methodology; Investigation; Data curation; Writing–review and editing. A.A.: Conceptualization; Supervision. W.F. and E.K. contributed equally.

## Generative AI disclosure

Generative AI tools (ChatGPT, OpenAI; Claude, Anthropic) assisted with code development and documentation, manuscript drafting and revision, reference checking and document preparation. Patient-level source data were not generated or altered by these tools. Numerical results were computed by the supplied analysis code. The authors are responsible for verifying AI-assisted content and the final submission.

## Data availability

Public TCGA-BRCA records are distributed through the NCI Genomic Data Commons (TCGA-BRCA project portal; Grossman et al., 2016) and UCSC Xena (Goldman et al., 2020). The accompanying code-and-data supplement includes the archived clinical and derived analysis inputs, input digests, exclusion ledger, analysis and simulation code, generated results and execution instructions. Reproduction starts from these supplied inputs; it does not reconstruct the earlier omics feature-selection pipeline from raw assay matrices. Only public, de-identified data are analyzed and no new participant contact occurs.

## Supplementary Material

Web Appendices A–J, Web Tables 1–12, Web Figures 1–5, and a code-and-data archive containing the derived analysis inputs, executable analysis and simulation code, generated numerical outputs, hashes, and execution instructions referenced in Sections 2–4 are available with this paper at the Biometrics website.

---

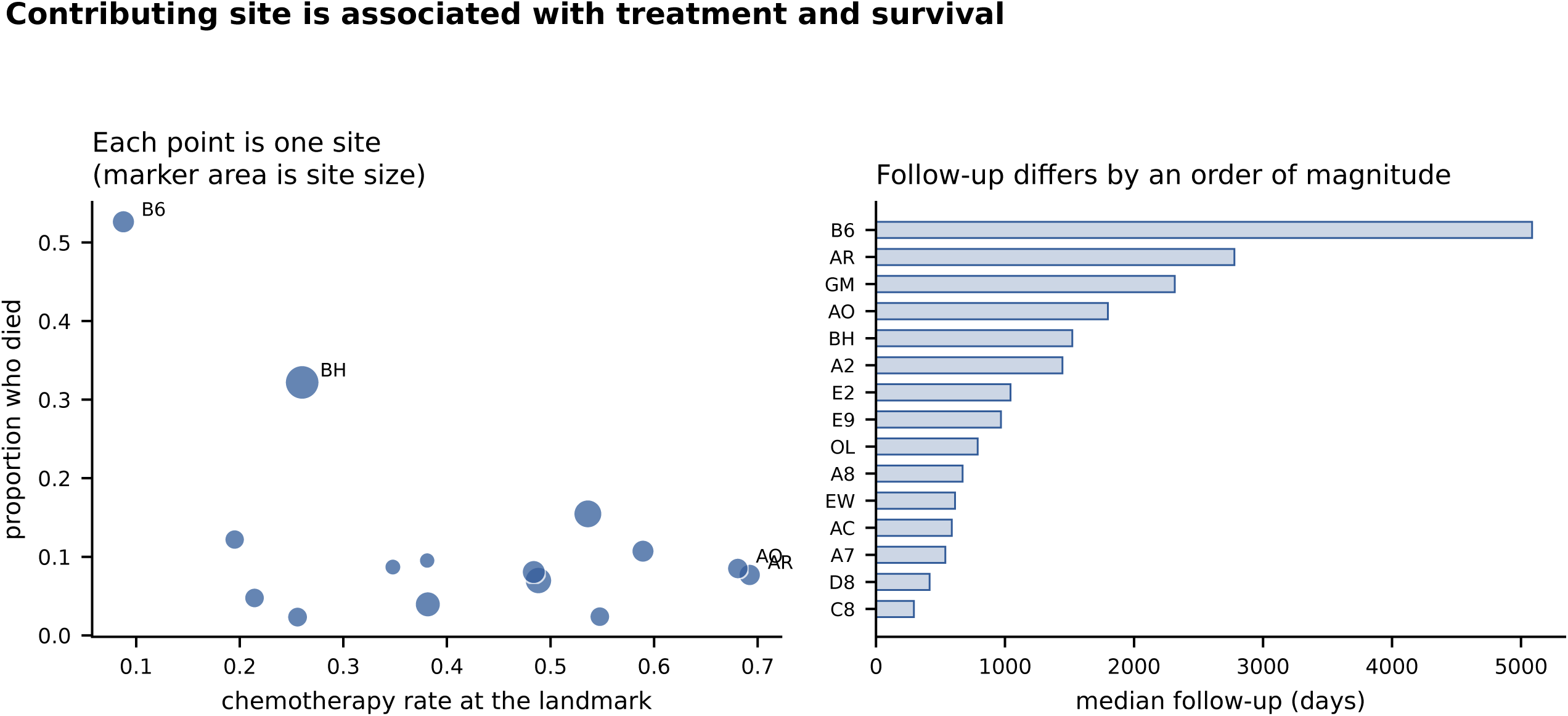

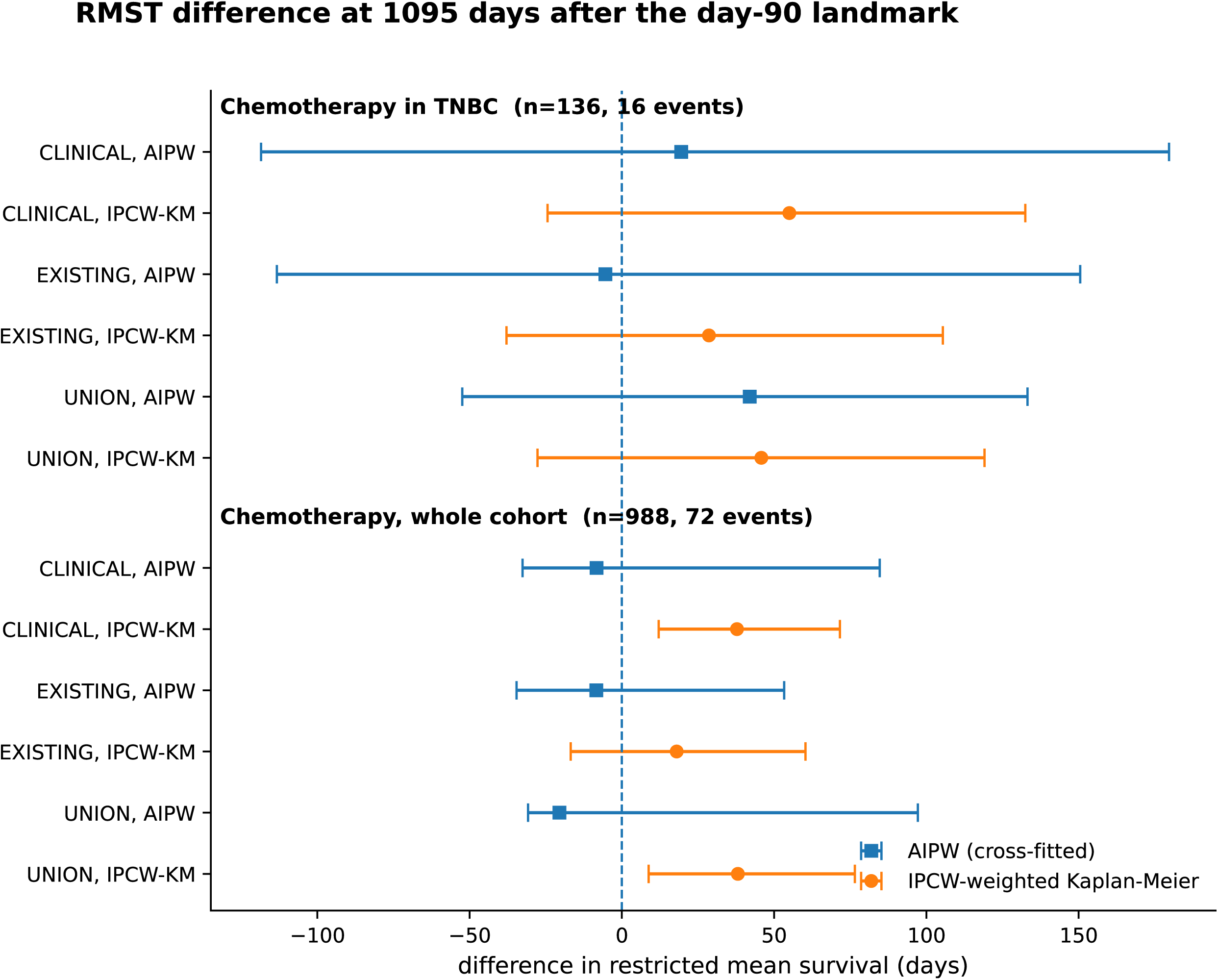

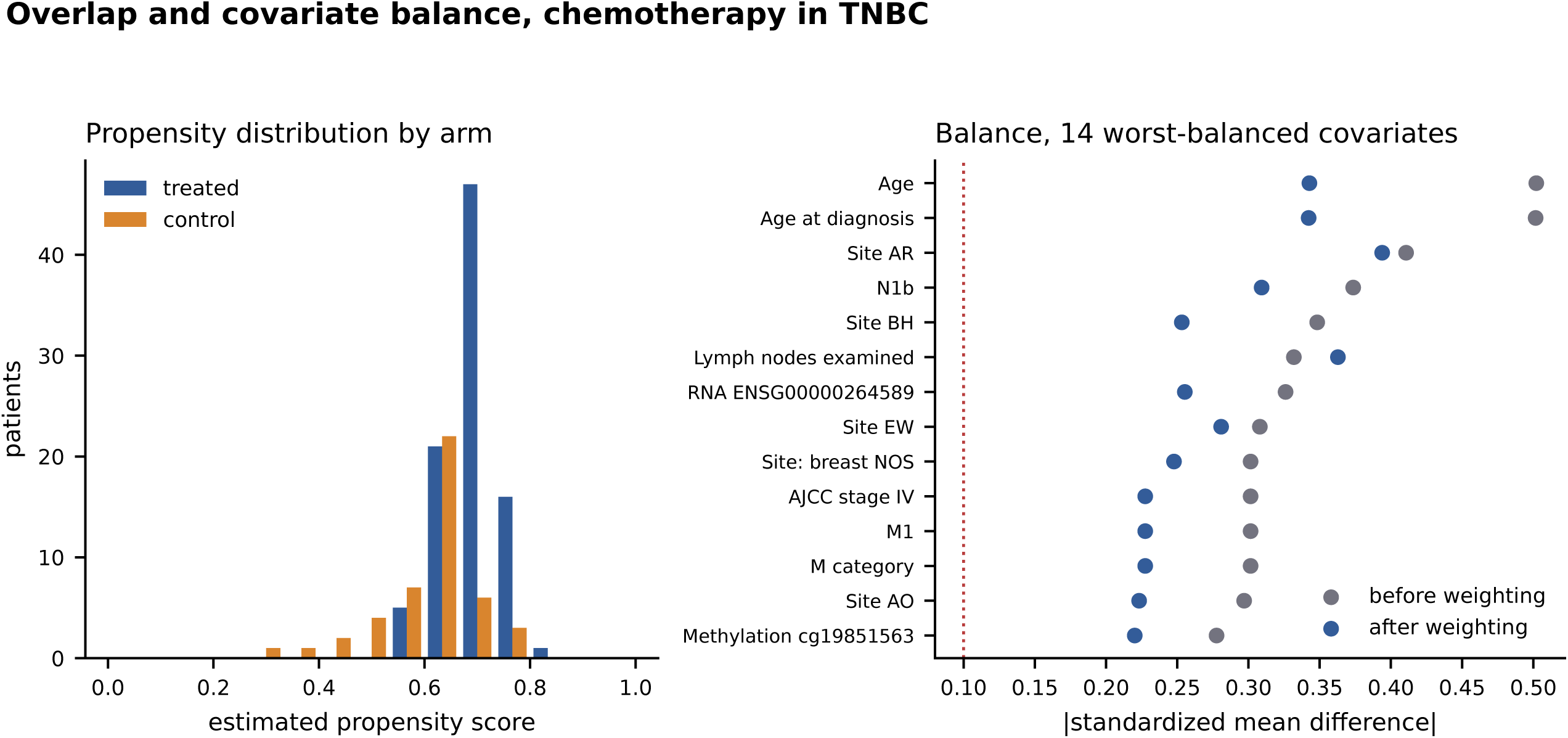

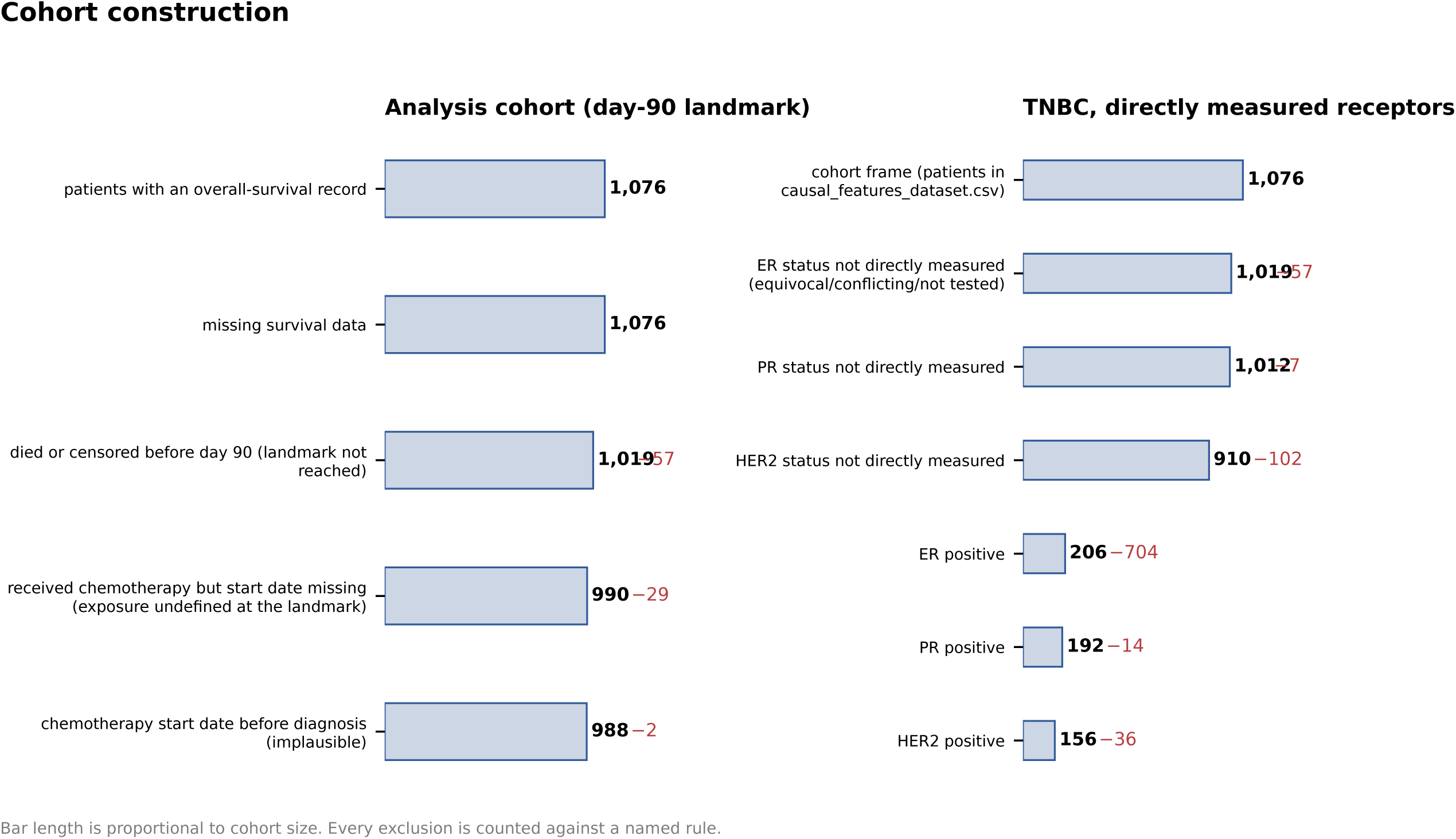

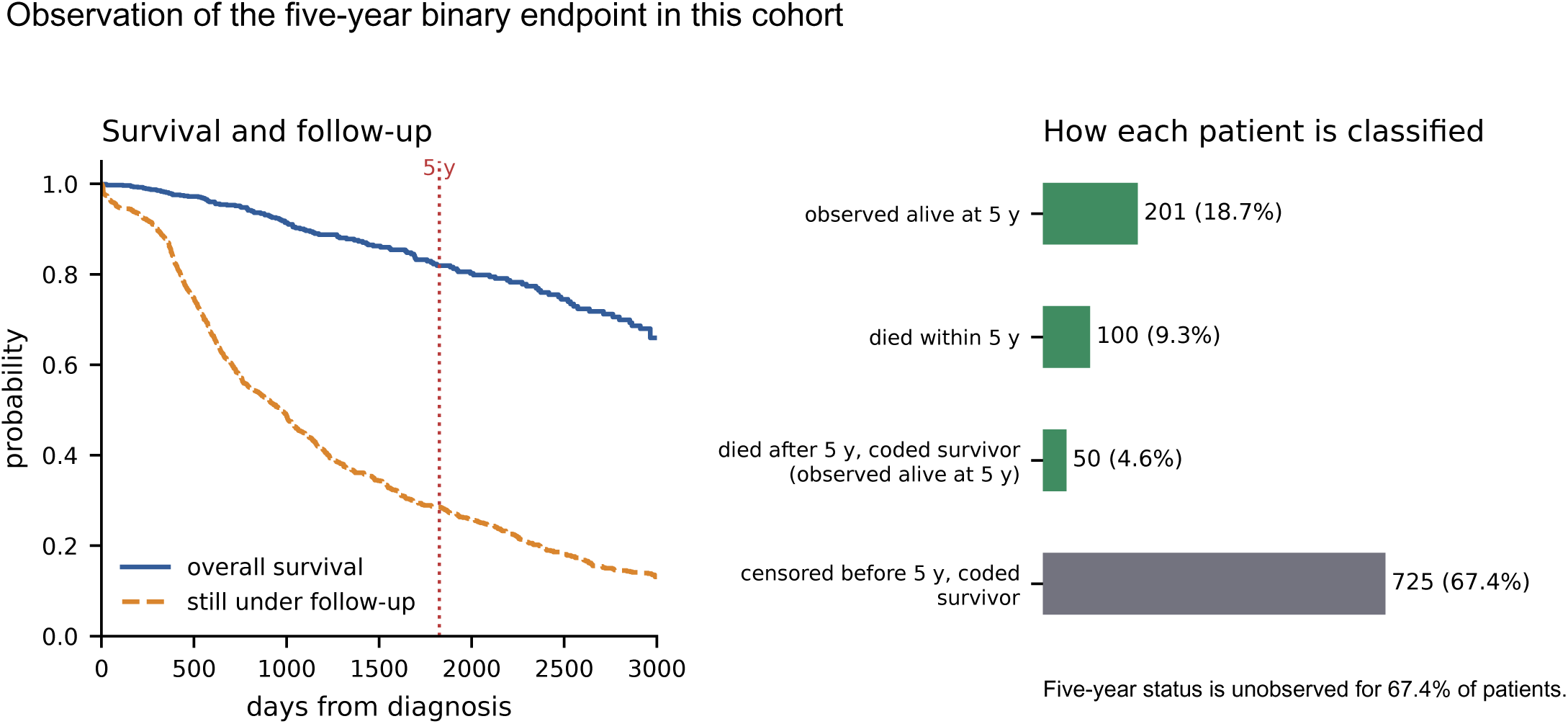

## Notes

### Competing Interest Statement

The authors have declared no competing interest.

### Author Declarations

Public TCGA-BRCA clinical and follow-up data analyzed in this study are available through the NCI Genomic Data Commons and UCSC Xena. The accompanying code-and-data supplement contains the archived clinical and derived analysis inputs, input digests, exclusion ledger, analysis and simulation code, generated results, and execution instructions. Only publicly available, de-identified human data were analyzed.

