## Supplementary for "Design and outcome-model estimability in doubly robust survival analysis: a breast cancer reanalysis"

This supplement accompanies the main article and provides the estimator equations, data audit, numerical results and reproducibility details. Clinical specification denotes the clinical adjustment set, covariate-conditional censoring and a 1095-day post-landmark horizon unless stated otherwise. The older reference validation and calibration are separated from the added Cox comparisons and independent stress scenarios.

### Web Appendix A. Estimators, assumptions and fixed preprocessing

All expectations in this appendix refer to an eligible landmark population. Let $R = \min(T, \tau)$, $Y = \min(\widetilde{T}, \tau)$, and $D$ indicate an observed death by $\tau$ or follow-up reaching $\tau$. Write $m_a^0(X) = E(R \mid A = a, X)$ and $e_0(X) = \Pr(A = 1 \mid X)$ for the true functions, and let $m_a(X)$ and $e(X)$ be working outcome and propensity functions. Correct censoring survival is denoted by $G$. Before stabilization, the complete-outcome AIPW contribution is

$$\begin{aligned}\psi(O) = {} & m_1(X) - m_0(X) \\ & + \frac{AD}{e(X)G(Y^- \mid 1, X)}\{Y - m_1(X)\} \\ & - \frac{(1-A)D}{\{1-e(X)\}G(Y^- \mid 0, X)}\{Y - m_0(X)\}.\end{aligned}$$

Under conditional independent censoring, $E(D \mid T, A, X) = G(R^- \mid A, X)$, with the stated tie convention. For correct $G$, conditioning on $A, X$ gives $E[D\{Y - m_a(X)\}/G(Y^- \mid a, X) \mid A = a, X] =$

$m_a^0(X) - m_a(X)$, where superscript 0 denotes the true conditional mean. Consequently,

$$\begin{aligned} E\{\psi(O) \mid X\} = m_1(X) - m_0(X) \\ + \frac{e_0(X)}{e(X)}\{m_1^0(X) - m_1(X)\} \\ - \frac{1-e_0(X)}{1-e(X)}\{m_0^0(X) - m_0(X)\}. \end{aligned}$$

This recovers $m_1^0 - m_0^0$ if $e = e_0$ or both outcome means are correct, conditional on correct $G$. It does not provide robustness to arbitrary misspecification of $G$, and fixed weight truncation need not retain this identity. It is not the full censoring-augmented efficient influence function.

Outcome regressions use complete cases weighted by $1/\widehat{G}$ inside treatment arms. The propensity is ridge logistic; the censoring process uses ridge Cox with treatment strata and a Breslow baseline. The unpenalized intercept is included in outcome/propensity parameter counts. The adaptive penalty is $\lambda = \max\{1, p\max(1, 10p/n)\}$ in the objective conventions in the code. For AIPW propensity and outcome regressions, $p, n$ refer to the full input matrix and sample; censoring fits compute their own training-sample penalty. Coefficients are trained on four of five folds. Folds are stratified jointly by treatment and observed death within the horizon.

Clinical stage and TNM are ordinally encoded. Numeric missing values use full-cohort medians with missing indicators. Sites with fewer than 20 patients in the landmark cohort are pooled; site coding is fixed before restricting to a subgroup. Receptors defining TNBC are never imputed. The archived multimodal matrix retains its previous processing; residual nonfinite entries are set to zero. These encodings and clinical imputations are fixed for resampling. Within an AIPW input matrix, propensity/outcome scaling also precedes fold splitting, whereas censoring-model scaling uses training observations. Bootstrap refits therefore condition on the constructed feature matrix and do not reproduce original feature discovery. No claim of fully nested model/feature selection is made.

Weighted KM clips propensity at 0.01/0.99; AIPW uses 0.02/0.98. Both floor $G$ at 0.05. AIPW caps positive combined weights at the pooled 99th percentile, clips outcome means to $[0, \tau]$ and clips the contrast to $[-\tau, \tau]$. The Cox outcome comparator instead has one unpenalized treatment coefficient, ridge-penalized standardized covariates, and a Breslow death-hazard baseline. Its arm-specific survival predictions are averaged over the same observed covariate distribution and integrated exactly between baseline-hazard knots. It does not use a separate censoring regression, relying on conditional independent censoring and correct specification of the outcome model. Treatment contrast and hazard ratio are distinct; only standardized RMST enters the comparison.

### Web Appendix B. Cohort audit and observed follow-up

The starting frame is the supplied earlier cohort, not every patient ever enrolled in TCGA-BRCA. Web Table 1 gives the sequential ledger: each exclusion count refers to the preceding row's survivors. Measured-receptor subgroup construction and original follow-up are shown in Web Figures 1–2. The treatment-field comparison establishes discordance, not an externally validated error rate.

### Web Appendix C. Complete effect estimates and proportional-hazards diagnostics

Web Table 2 reports the original adjustment-set and horizon sensitivity estimates. The unadjusted KM and Cox clinical benchmarks and their bootstrap intervals are in main Table 2. They use 2000 new patient bootstrap draws, paired with each other. Earlier KM intervals for a correction ladder rung use their original seed, so Monte Carlo differences in endpoints do not imply different point estimates. The complete proportional-hazards screens appear later in Web Table 11.

**Web Appendix D. Censoring, horizon support and five-device stabilization**

Censoring diagnostics are evaluated at each complete case's restricted time. They differ between AIPW's training-fold censoring fits and the primary estimator's full-sample fit. The combined-weight ESS values reported in the article refer to capped AIPW complete-outcome residual weights; they are not an effective sample size for the time-varying KM risk sets. Selected-horizon risk counts and estimator weight diagnostics are supplied as CSV files. Sites with short follow-up motivate, but do not validate, conditional censoring models.

**Web Appendix E. Landmark, fold information and seed stability**

Changing to day 180 changes both eligibility and exposure status. It is a sensitivity analysis for a different landmark population, not a bound on selection bias. The outcome effective degrees of freedom are computed from the actual training-fit matrix and Cox-derived weights as $\mathrm{tr}\{D(D^\top WD+P)^{-1}D^\top W\}$, where the intercept entry of $P$ is zero. Fold-specific complete counts, dimension, penalty and effective degrees of freedom are supplied in `20_outcome_fold_df.csv`. Nominal rho counts use complete outcomes, not deaths alone.

**Web Appendix F. Original diagnostic calibration**

The 32-cell calibration uses four sample sizes (150, 300, 600, 1000), four covariate dimensions (10, 25, 50, 75) and two baseline event-rate settings (0.12, 0.30). Each cell has 60 replicates. Covariates are independent standard Gaussian. Treatment follows a logistic model with coefficients $(0.7,-0.5,0.3)$ on the first three covariates and a centered linear predictor plus log odds for 0.6. Survival is exponential with hazard $b\exp(0.5X_1+0.3X_2)0.65^A$, where $b=-\log(1-r)/1095$. Censoring is exponential with mean $1.4\times 1095$ and independent of covariates. Let $d$ denote the input covariate dimension, before adding an intercept. Adjustment sets are nested prefixes of size $\max(3,\lfloor d/4\rfloor)$, $\max(4,\lfloor d/2\rfloor)$ and $d$. All sets contain the assignment predictors. For estimator $m$, cell $c$ and replicate $r$, let $\widehat{\Delta}_{mcrs}$ be the estimate under nested adjustment set $s\in\{1,2,3\}$, with $s=3$ the full set. With $R_c$ retained replicates, the calibration response is

$$Q_{mc}=\frac{R_c^{-1}\sum_{r=1}^{R_c}\left\{\max_s\widehat{\Delta}_{mcrs}-\min_s\widehat{\Delta}_{mcrs}\right\}}{\mathrm{SD}_{r=1,\ldots,R_c}(\widehat{\Delta}_{mcr3})}. \tag{S1}$$

The denominator is the sample standard deviation across full-set replicate estimates (divisor $R_c-1$), not a within-replicate standard deviation or a bootstrap standard error. Thus the range is computed first within each replicate, then averaged, and only then divided by the full-set sampling standard deviation. Replicates with fewer than 15 patients in either treatment group are omitted by the original grid generator. When computing $\rho$, the grid implementation uses $\max\{c_{\min}(1-1/K),1\}$ in its denominator to avoid division by a very small complete-outcome count. The reported $\rho$ is averaged over replicates in a cell. The fitted response is $Q_{mc}=b_0+b_1\log(\overline{\rho}_c)$, and the warning crossing at level $q$ is $\exp\{(q-b_0)/b_1\}$. The log-linear fit and its crossing use 2000 bootstrap resamples of grid cells. These intervals reflect variation across the selected cells and fit, not uncertainty over all possible data-generating regimes. They also do not propagate every source of within-cell Monte Carlo uncertainty. Penalty scales 0.3 and 3 use 30 replicates per cell; the specified scale uses 60. The nested sets do not establish a lower bound on sensitivity under arbitrary real adjustment sets.

#### Web Appendix G. Validation against known truth and independent stress scenarios

The original operating-point validation uses 500 replicates per scenario and 500 inner patient bootstrap replicates. It simulates Gaussian covariates and exponential survival, logistic treatment and censoring depending on a survival predictor. The input covariate counts are 25 and 28 before adding an intercept; these approximate, rather than exactly match, the clinical outcome-model dimensions. Its event targets and treated fractions are application-motivated. Empirical coverage cannot be assigned to the clinical intervals merely because sample size and event counts are similar.

The independent stress study has six cells: $n = 136, p = 24$ or $n = 988, p = 27$, crossed with three mechanisms. There are 200 replicates and 199 inner patient bootstraps per cell. New simulation seeds are distinct from the original calibration. Gaussian scenarios use independent standard-normal covariates; the nonlinear scenario uses independent $t_3/\sqrt{3}$ covariates. The default propensity is $\mathrm{expit}(0.7X_1 - 0.5X_2 + 0.3X_3)$. The control death hazard is $h_0 = b\exp(0.5X_1 + 0.3X_2)$, and the censoring hazard is $(\log 2/1100)\exp(0.5X_1 + 0.5X_3)$. Thus censoring depends on measured survival predictors. The PH treatment hazard ratio is 0.65. The delayed-benefit scenario instead uses treatment hazard ratio 1.2 through day 365 and 0.45 thereafter; it violates a constant treatment hazard ratio in the Cox outcome model.

In the nonlinear heavy-tail scenario, add $0.61\mathbf{1}(X_1 > 1)$ to the propensity linear predictor, replace the death predictor by $0.5X_1 + 0.5\sin(X_2) + 0.3(X_3^2 - 1)$, and add $0.35(X_2^2 - 1)$ to the censoring predictor. Linear nuisance regressions are then misspecified. To keep the generated hazards finite, death/censoring predictors are clipped to $[-4, 4]$ and the propensity predictor to $[-20, 20]$. The baseline $b$ is calibrated by deterministic-seed numerical averaging over 30000 reference covariates to expected observed deaths near 16 or 72. The exact fitted baseline constants are saved in `21_stress_design.json`; achieved mean deaths are in the replicate and summary CSV files.

For hazards $h_1$ before $u = 365$ and $h_2$ afterward, the restricted mean is $(1 - e^{-h_1 u})/h_1 + e^{-h_1 u}(1 - e^{-h_2(\tau - u)})/h_2$. The treated-minus-control difference is averaged over each replicate's covariates to obtain its exact conditional standardized truth. Four estimators are compared against this truth; the unadjusted KM benchmark is expected to retain confounding. All estimators use their stated penalties and stabilization. The AIPW fold seed is fixed within a replicate's bootstrap procedure. A replicate is evaluable for a method only if its point estimate is finite and at least 95% of its inner fits succeed. Failures are reported; coverage is conditional on evaluability. Bias Monte Carlo SE is the SD of estimation errors divided by the square root of evaluable replicates. Wilson intervals describe binomial Monte Carlo uncertainty in coverage. RMSE is relative to each replicate's conditional truth. The full table also reports unadjusted KM; the main coverage figure shows the three adjusted methods.

Schoenfeld rank-time screens use lifelines Efron-tie reference refits for the death and censoring processes. Penalties are converted to the code's scale, the treatment coefficient is unpenalized in the death model, and censoring is stratified by treatment. Holm adjustment is within each model over covariates. These exploratory checks use penalized fits, are not exact finite-sample tests, and cannot verify independent censoring. They are reported as potential model-fit concerns, not automatic model acceptance/rejection.

### Web Appendix H. Exploratory heterogeneity and scope of validation

The earlier analysis gated heterogeneity at 50 deaths, a convention rather than a guarantee of usable power. Only whole-cohort estimates are retained here. Their AIPW scores are cross-fitted for nuisance estimation, but the later linear ranking was fitted and evaluated on the same cohort. The old subgroup, BLP and RATE intervals resampled already computed scores rather than the full estimation procedure. They are therefore omitted from this submission's heterogeneity table. This avoids presenting conditional fixed-score intervals as if they reflected all nuisance, ranking and selection uncertainty. The following estimates are descriptive only and do not establish targeting ability or effect modification. Confirmatory RATE evaluation (Yadlowsky et al., 2025) would require an independently learned ranking or an appropriate additional cross-fitting scheme; nominal significance of the old interval is not used as evidence.

For context, the earlier approximate minimum-detectable subgroup differences are TNBC: 279.2 days; Whole cohort: 82.2 days. These use $2(z_{0.975}+z_{0.80})s/\sqrt{nq(1-q)}$, where $q$ is the treated fraction and $s$ is the restricted-time SD among complete cases. They are rough complete-data calculations, not power calculations for the weighted survival estimator, censoring or a pre-specified interaction test. The 50-death gate and this approximation do not establish adequate power.

Classification and clustering metrics do not evaluate the RMST contrast. The molecular variables in the inherited adjustment representation were not selected in this study as a biological signature. GO/KEGG or other gene-set enrichment would require a defined gene universe and molecular-selection target and, even if performed, would characterize the provenance of selected covariates rather than validate the RMST treatment contrast or its identifying assumptions. Statistical validation instead examines the specified estimators against known RMST truth, alongside support, fit and sensitivity diagnostics. The related prognostic study addresses transport and incremental predictive utility separately.

### Web Appendix I. Reproducibility, software and files

The code-and-data supplement provides the clinical and derived inputs, generated CSV outputs, software requirements, seed and source hashes, and a single pipeline entry point. The analysis uses NumPy/Pandas; figures use Matplotlib, and the comparator cross-checks and PH screens use lifelines/SciPy. LaTeX compilation is included in the documented build command. The original Kaplan–Meier, RMST and Cox-censoring cross-checks are retained in `15_lifelines_crosscheck.csv`. The new Cox RMST implementation is independently checked against lifelines on untied unpenalized data, under treatment relabeling, constant-column addition and feasible-range constraints. Agreement of a numerical component does not validate a clinical model.

The supplied raw clinical exports and derived survival/feature tables suffice for the analyses in this paper. The original raw omics matrices and Xena source export are not reconstructed. Input SHA-256 digests and the distinction between supplied inputs and upstream provenance are documented in the plain-text README. The bootstrap and simulation replication counts are recorded in their output tables. Reduced smoke runs write into a separate directory and are never used to generate the submission PDFs.

### Web Appendix J. Related work, archived implementation and scope

The earlier covariate list was intended as an input to a published feature-selection framework's downstream analysis. The archived implementation accepted a feature-list argument but substituted a univariate penalized-Cox screen. The source location is in the feature-extraction tree. This observation concerns that archived routine; it does not establish which routine generated the published framework's results, and no claim is made that those published results are affected or unaffected. The supplied source snapshot and audit provenance identify the limited implementation claim. The feature-selection framework itself is not re-evaluated in the current manuscript.

The related sequencing manuscript (Krikun and Alkhateeb, 2026a) studies hormone-therapy initiation by day 180 in verified HR-positive, HER2-negative TCGA-BRCA. It examines upstream chemotherapy timing using an outcome-blind sequencing audit and compares full-cohort adjustment with sequencing-aware restriction. Its target is an overlap-population contrast over 730 days, rather than the day-90 chemotherapy contrast over 1095 days examined here. Its sequencing-aware clinical population has 271 patients and 36 deaths. TCGA data, treatment-record reconstruction and RMST methods are shared; the treatment question, eligibility rule, target population and simulation objective differ.

The TCGA–METABRIC manuscript (Krikun and Alkhateeb, 2026c) evaluates assayability, fixed-panel prognostic transport and incremental predictive utility, with a separate nested reconstruction of the feature-selection framework in METABRIC. It shares TCGA source data and earlier feature-selection context with this study, but does not evaluate the present chemotherapy contrast. Neither related manuscript is independent clinical validation of the current analysis. The published Markov-blanket framework (Krikun and Alkhateeb, 2026b) is cited for provenance, not re-evaluated here.

Web Table 1: Day-90 analysis cohort ledger.

| Rule | Excluded | Remaining |
| --- | --- | --- |
| patients with an overall-survival record | – | 1076 |
| missing survival data | 0 | 1076 |
| died or censored before day 90 (landmark not reached) | 57 | 1019 |
| received chemotherapy but start date missing (exposure undefined at the landmark) | 29 | 990 |
| chemotherapy start date before diagnosis (implausible) | 2 | 988 |

Web Table 2: All original adjusted RMST estimates. Intervals are patient-bootstrap percentile intervals.

| Population/set | Horizon | Estimator | RMST (95% interval) |
|---|---|---|---|
| TNBC / Clinical | 1095 | AIPW | 19.5 (-118.5, 179.7) |
| TNBC / Clinical | 1095 | Weighted KM | 55.0 (-24.4, 132.5) |
| TNBC / Existing | 1095 | AIPW | -5.4 (-113.3, 150.5) |
| TNBC / Existing | 1095 | Weighted KM | 28.6 (-37.9, 105.4) |
| TNBC / Union | 1095 | AIPW | 42.0 (-52.4, 133.2) |
| TNBC / Union | 1095 | Weighted KM | 45.8 (-27.7, 119.1) |
| TNBC / Clinical | 1825 | AIPW | 383.8 (107.5, 670.3) |
| TNBC / Clinical | 1825 | Weighted KM | 256.2 (31.9, 515.5) |
| TNBC / Existing | 1825 | AIPW | 13.0 (-449.3, 596.4) |
| TNBC / Existing | 1825 | Weighted KM | 121.1 (-74.9, 359.3) |
| TNBC / Union | 1825 | AIPW | 333.3 (35.5, 602.3) |
| TNBC / Union | 1825 | Weighted KM | 226.7 (9.5, 468.0) |
| Whole cohort / Clinical | 1095 | AIPW | -8.3 (-32.6, 84.7) |
| Whole cohort / Clinical | 1095 | Weighted KM | 37.8 (12.1, 71.6) |
| Whole cohort / Existing | 1095 | AIPW | -8.4 (-34.6, 53.3) |
| Whole cohort / Existing | 1095 | Weighted KM | 18.0 (-16.8, 60.3) |
| Whole cohort / Union | 1095 | AIPW | -20.5 (-30.8, 97.2) |
| Whole cohort / Union | 1095 | Weighted KM | 38.1 (8.8, 76.5) |
| Whole cohort / Clinical | 1825 | AIPW | 151.1 (34.7, 353.8) |
| Whole cohort / Clinical | 1825 | Weighted KM | 119.1 (44.8, 208.8) |

| Population/set | Horizon | Estimator | RMST (95% interval) |
|---|---|---|---|
| Whole cohort / Existing | 1825 | AIPW | -42.7 (-130.7, 149.5) |
| Whole cohort / Existing | 1825 | Weighted KM | 108.7 (22.6, 226.6) |
| Whole cohort / Union | 1825 | AIPW | 134.3 (52.9, 350.4) |
| Whole cohort / Union | 1825 | Weighted KM | 124.5 (49.0, 219.1) |

Web Table 3: Estimator-specific balance. Values are absolute standardized mean differences (SMD). Treatment weights alone assess baseline balance; complete-outcome censoring weights define a different diagnostic.

| Population | Weight definition | Mean SMD | Max SMD | No. $> 0.1$ |
|---|---|---|---|---|
| TNBC | unweighted | 0.173 | 0.502 | 16 |
| TNBC | primary KM treatment weights | 0.113 | 0.379 | 11 |
| TNBC | AIPW treatment weights | 0.130 | 0.380 | 14 |
| Whole cohort | unweighted | 0.173 | 0.632 | 19 |
| Whole cohort | primary KM treatment weights | 0.051 | 0.196 | 4 |
| Whole cohort | AIPW treatment weights | 0.049 | 0.216 | 3 |

Web Table 4: Original censoring-model sensitivity. Floor counts are complete cases with fitted censoring survival below 0.05.

| Population/method | Model | RMST (95% interval) | $G_{\min}$ | Floor |
|---|---|---|---|---|
| TNBC / AIPW | Cox | 19.5 (-118.5, 179.7) | 0.027 | 2 |
| TNBC / AIPW | Arm KM | 75.5 (-42.6, 170.1) | 0.324 | 0 |
| TNBC / Weighted KM | Cox | 55.0 (-24.4, 132.5) | 0.078 | 0 |
| TNBC / Weighted KM | Arm KM | 52.1 (-27.7, 132.6) | 0.386 | 0 |
| Whole cohort / AIPW | Cox | -8.3 (-32.6, 84.7) | 0.002 | 7 |
| Whole cohort / AIPW | Arm KM | 28.4 (-15.5, 86.1) | 0.412 | 0 |
| Whole cohort / Weighted KM | Cox | 37.8 (12.1, 71.6) | 0.007 | 4 |
| Whole cohort / Weighted KM | Arm KM | 32.6 (10.2, 57.7) | 0.418 | 0 |

Web Table 5: Updated stabilizer sensitivity, in RMST days. The joint relaxation now disables both prediction and contrast clipping, sets the propensity limits to 0.001/0.999 and censoring floor to 0.01, and removes the combined-weight cap. These remain near-untruncated rather than literally unbounded weights.

| Population | Variant | AIPW | Weighted KM |
|---|---|---|---|
| TNBC | specified | 19.5 | 55.0 |
| TNBC | propensity clip 0.01/0.99 | 19.5 | 55.0 |
| TNBC | propensity clip 0.001/0.999 | 19.5 | 55.0 |
| TNBC | censoring floor 0.02 | -52.0 | 55.0 |
| TNBC | censoring floor 0.10 | 69.4 | 55.6 |
| TNBC | combined weight cap 0.999 | 10.9 | 55.0 |
| TNBC | combined weight cap 0.95 | 100.2 | 55.0 |
| TNBC | outcome prediction clipping off | 19.8 | 55.0 |
| TNBC | contrast clipping off | 19.5 | 55.0 |
| TNBC | all five devices relaxed | -53.0 | 55.0 |
| TNBC | arm-only censoring | 75.5 | 52.1 |
| Whole cohort | specified | -8.3 | 37.8 |
| Whole cohort | propensity clip 0.01/0.99 | -8.3 | 37.8 |
| Whole cohort | propensity clip 0.001/0.999 | -8.3 | 37.8 |
| Whole cohort | censoring floor 0.02 | -12.2 | 42.9 |
| Whole cohort | censoring floor 0.10 | 13.2 | 36.4 |

| Population | Variant | AIPW | Weighted KM |
|---|---|---|---|
| Whole cohort | combined weight cap 0.999 | -4.7 | 37.8 |
| Whole cohort | combined weight cap 0.95 | 19.4 | 37.8 |
| Whole cohort | outcome prediction clipping off | -9.1 | 37.8 |
| Whole cohort | contrast clipping off | -8.3 | 37.8 |
| Whole cohort | all five devices relaxed | -19.7 | 48.0 |
| Whole cohort | arm-only censoring | 28.4 | 32.6 |

Web Table 6: Day-180 landmark sensitivity at a 1095-day horizon.

| Population | *n*/deaths | Estimator | RMST (95% interval) |
|---|---|---|---|
| TNBC | 133/16 | AIPW | 55.0 (-156.4, 277.2) |
| TNBC | 133/16 | Weighted KM | 83.3 (-40.6, 201.6) |
| Whole cohort | 971/69 | AIPW | 26.5 (-6.2, 123.1) |
| Whole cohort | 971/69 | Weighted KM | 44.5 (14.0, 78.7) |

Web Table 7: Original six-seed repeat. This separates computational reproducibility from partition and simulation Monte Carlo variability.

| Quantity | MC SD | Mean interval width | Interval reading |
|---|---|---|---|
| TNBC IPCW-KM | 0.00 | 156.60 | interval spans zero |
| TNBC AIPW | 24.83 | 291.55 | interval spans zero |
| Whole cohort IPCW-KM | 0.00 | 59.89 | interval excludes zero |
| Whole cohort AIPW | 14.86 | 115.06 | interval spans zero |
| rho threshold (calibration) | 0.02 | 0.07 | triple-negative above, whole cohort below |

Web Table 8: Original warning-level sensitivities. The specified crossing is 0.50; the threshold is not an estimator-validity criterion.

| Varied | Value | Threshold | 95% cell-bootstrap |
|---|---|---|---|
| Crossing | 0.25 | 0.11 | 0.09, 0.14 |
| Crossing | 0.50 | 0.31 | 0.28, 0.35 |
| Crossing | 0.75 | 0.88 | 0.74, 1.10 |
| Crossing | 1.00 | 2.47 | 1.84, 3.70 |
| Penalty multiplier | 0.3 | 0.22 | 0.19, 0.27 |
| Penalty multiplier | 1.0 | 0.31 | 0.28, 0.35 |
| Penalty multiplier | 3.0 | 1.32 | 0.70, 4.94 |

Web Table 9: Original operating-point validation. SD is empirical estimate SD; SE is mean bootstrap SE. Coverage refers only to these simulated scenarios.

| Scenario/method | Truth | Bias | SD/SE | Coverage |
|---|---|---|---|---|
| TNBC-like / AIPW | 44.2 | -1.4 | 65.1/63.4 | 95.0% |
| TNBC-like / Weighted KM | 44.2 | -12.5 | 54.5/50.7 | 92.2% |
| cohort-like / AIPW | 24.2 | 1.1 | 17.2/18.7 | 95.8% |
| cohort-like / Weighted KM | 24.2 | -3.0 | 15.4/15.8 | 94.0% |

Web Table 10: Independent stress validation. Bias is accompanied by its Monte Carlo SE. Coverage (percent) includes a 95% Wilson Monte Carlo interval. Failures are out of 200 replicates per cell and estimator. All individual results are supplied.

| Method | Bias (MC SE) | RMSE | Coverage (MC interval) | Failed |
|---|---|---|---|---|
| **PH / conditional censoring;** $n = 136$ | | | | |
| AIPW | 3.4 (3.6) | 50.8 | 97.5 (94.3, 98.9) | 0 |
| Cox RMST | -13.3 (2.6) | 39.1 | 91.5 (86.8, 94.6) | 0 |
| Weighted KM | -14.0 (3.1) | 46.5 | 93.0 (88.6, 95.8) | 0 |
| Unadjusted KM | -19.5 (3.1) | 48.3 | 95.0 (91.0, 97.3) | 0 |
| **PH / conditional censoring;** $n = 988$ | | | | |
| AIPW | 5.2 (1.5) | 21.5 | 94.0 (89.8, 96.5) | 0 |
| Cox RMST | 0.2 (0.9) | 12.5 | 92.0 (87.4, 95.0) | 0 |
| Weighted KM | -0.7 (1.3) | 18.6 | 91.5 (86.8, 94.6) | 0 |
| Unadjusted KM | -8.6 (1.0) | 17.1 | 85.5 (80.0, 89.7) | 0 |
| **Delayed benefit;** $n = 136$ | | | | |
| AIPW | 6.2 (4.2) | 59.8 | 97.0 (93.6, 98.6) | 0 |
| Cox RMST | 14.7 (2.7) | 40.7 | 91.0 (86.2, 94.2) | 1 |
| Weighted KM | -13.5 (3.5) | 50.8 | 92.5 (88.0, 95.4) | 0 |
| Unadjusted KM | -16.4 (3.4) | 50.8 | 93.5 (89.2, 96.2) | 0 |

| Method | Bias (MC SE) | RMSE | Coverage (MC interval) | Failed |
|---|---|---|---|---|
| **Delayed benefit;** $n = 988$ | | | | |
| AIPW | 4.8 (1.5) | 21.0 | 93.0 (88.6, 95.8) | 0 |
| Cox RMST | 19.2 (0.8) | 22.5 | 63.5 (56.6, 69.9) | 0 |
| Weighted KM | -0.7 (1.3) | 18.1 | 92.0 (87.4, 95.0) | 0 |
| Unadjusted KM | -8.2 (1.1) | 17.3 | 90.5 (85.6, 93.8) | 0 |
| **Nonlinear / heavy tail;** $n = 136$ | | | | |
| AIPW | 1.2 (3.7) | 51.7 | 98.5 (95.7, 99.5) | 0 |
| Cox RMST | -5.7 (2.5) | 36.1 | 93.0 (88.6, 95.8) | 0 |
| Weighted KM | -5.5 (3.2) | 46.1 | 94.5 (90.4, 96.9) | 0 |
| Unadjusted KM | -5.3 (3.2) | 45.0 | 95.5 (91.7, 97.6) | 0 |
| **Nonlinear / heavy tail;** $n = 988$ | | | | |
| AIPW | -2.6 (1.3) | 18.9 | 93.0 (88.6, 95.8) | 0 |
| Cox RMST | -0.4 (0.8) | 11.0 | 92.5 (88.0, 95.4) | 0 |
| Weighted KM | -3.5 (1.4) | 20.3 | 89.5 (84.5, 93.0) | 0 |
| Unadjusted KM | -5.7 (1.1) | 16.5 | 88.5 (83.3, 92.2) | 0 |

Web Table 11: Complete proportional-hazards screens. Raw and within-model Holm-adjusted $p$-values from Efron reference refits. A dash indicates a term not included in that process model. Tests are exploratory; failure to reject does not establish proportional hazards.

| | Death model | | Censoring model | |
|---|---|---|---|---|
| Covariate | Raw $p$ | Holm $p$ | Raw $p$ | Holm $p$ |
| **TNBC** | | | | |
| age | 0.2747 | 1 | 0.5401 | 1 |
| Chemotherapy by landmark | 0.215 | 1 | – | – |
| m_stage | 0.9013 | 1 | 0.4205 | 1 |
| m_stage_missing | 0.3966 | 1 | 0.5916 | 1 |
| n_stage | 0.1174 | 1 | 0.6095 | 1 |
| site_A2 | 0.9742 | 1 | 0.8813 | 1 |
| site_A7 | 0.8577 | 1 | 0.9195 | 1 |
| site_A8 | 0.7965 | 1 | 0.679 | 1 |
| site_AC | 0.7188 | 1 | 0.7895 | 1 |
| site_AO | 0.9912 | 1 | 0.3824 | 1 |
| site_AR | 0.8034 | 1 | 0.3135 | 1 |
| site_B6 | 0.633 | 1 | 0.3775 | 1 |
| site_BH | 0.9795 | 1 | 0.8489 | 1 |
| site_C8 | 0.6166 | 1 | 0.9864 | 1 |
| site_D8 | 0.8823 | 1 | 0.7598 | 1 |

| | Death model | | Censoring model | |
|---|---|---|---|---|
| Covariate | Raw *p* | Holm *p* | Raw *p* | Holm *p* |
| site_E2 | 0.8408 | 1 | 0.6786 | 1 |
| site_E9 | 0.9545 | 1 | 0.7904 | 1 |
| site_EW | 0.7372 | 1 | 0.7165 | 1 |
| site_GM | 0.721 | 1 | 0.4778 | 1 |
| site_OL | 0.7565 | 1 | 0.9207 | 1 |
| site_OTHER | 0.9205 | 1 | 0.9551 | 1 |
| stage | 0.3674 | 1 | 0.8732 | 1 |
| stage_missing | 0.8821 | 1 | 0.6672 | 1 |
| t_stage | 0.4404 | 1 | 0.7585 | 1 |
| year_dx | 0.4579 | 1 | 0.9507 | 1 |
| **Whole cohort** | | | | |
| age | 0.2479 | 1 | 0.3094 | 1 |
| Chemotherapy by landmark | 0.9621 | 1 | – | – |
| m_stage | 0.7122 | 1 | 0.8586 | 1 |
| m_stage_missing | 0.2252 | 1 | 0.04202 | 1 |
| n_stage | 0.8597 | 1 | 0.9458 | 1 |
| n_stage_missing | 0.9364 | 1 | 0.599 | 1 |
| site_A2 | 0.8134 | 1 | 0.8195 | 1 |
| site_A7 | 0.7927 | 1 | 0.5087 | 1 |
| site_A8 | 0.3653 | 1 | 0.1479 | 1 |
| site_AC | 0.4389 | 1 | 0.3223 | 1 |
| site_AO | 0.3319 | 1 | 0.1056 | 1 |

| | Death model | | Censoring model | |
|---|---|---|---|---|
| Covariate | Raw *p* | Holm *p* | Raw *p* | Holm *p* |
| site_AR | 0.09758 | 1 | 0.6939 | 1 |
| site_B6 | 0.2263 | 1 | 0.9485 | 1 |
| site_BH | 0.3245 | 1 | 0.4581 | 1 |
| site_C8 | 0.5864 | 1 | 0.4829 | 1 |
| site_D8 | 0.9438 | 1 | 0.637 | 1 |
| site_E2 | 0.7668 | 1 | 0.9571 | 1 |
| site_E9 | 0.2856 | 1 | 0.966 | 1 |
| site_EW | 0.8112 | 1 | 0.6094 | 1 |
| site_GM | 0.3529 | 1 | 0.6969 | 1 |
| site_OL | 0.6205 | 1 | 0.9758 | 1 |
| site_OTHER | 0.2142 | 1 | 0.01529 | 0.3975 |
| stage | 0.4257 | 1 | 0.9606 | 1 |
| stage_missing | 0.6634 | 1 | 0.7019 | 1 |
| t_stage | 0.1907 | 1 | 0.8791 | 1 |
| t_stage_missing | 0.7374 | 1 | 0.7006 | 1 |
| year_dx | 0.08829 | 1 | 8.414e-05 | 0.002272 |
| year_dx_missing | 0.1955 | 1 | 0.7684 | 1 |

Web Table 12: Whole-cohort descriptive heterogeneity summaries. No confirmatory intervals are assigned. BLP denotes best linear projection; AUTOC is an in-sample ranking summary.

| Analysis | Term | $n$ | Estimate, days |
|---|---|---|---|
| group effect | age $< 50$ | 272 | -8.6 |
| group effect | age $\geq 50$ | 716 | -8.2 |
| group effect | node negative (N0) | 467 | -31.2 |
| group effect | node positive (N1-3) | 506 | 46.5 |
| group effect | stage I-II | 733 | -31.2 |
| group effect | stage III-IV | 233 | 66.8 |
| BLP of CATE | intercept (ATE) | 988 | -8.3 |
| BLP of CATE | age (centred) | 988 | 2.1 |
| BLP of CATE | node positive | 988 | 100.7 |
| BLP of CATE | stage III-IV | 988 | 40.8 |
| CATE calibration | quintile 1 | 198 | -6.6 |
| CATE calibration | quintile 2 | 200 | -84.9 |
| CATE calibration | quintile 3 | 199 | -65.3 |
| CATE calibration | quintile 4 | 203 | -31.2 |
| CATE calibration | quintile 5 | 188 | 156.3 |
| RATE (AUTOC) | AUTOC | 988 | 79.4 |

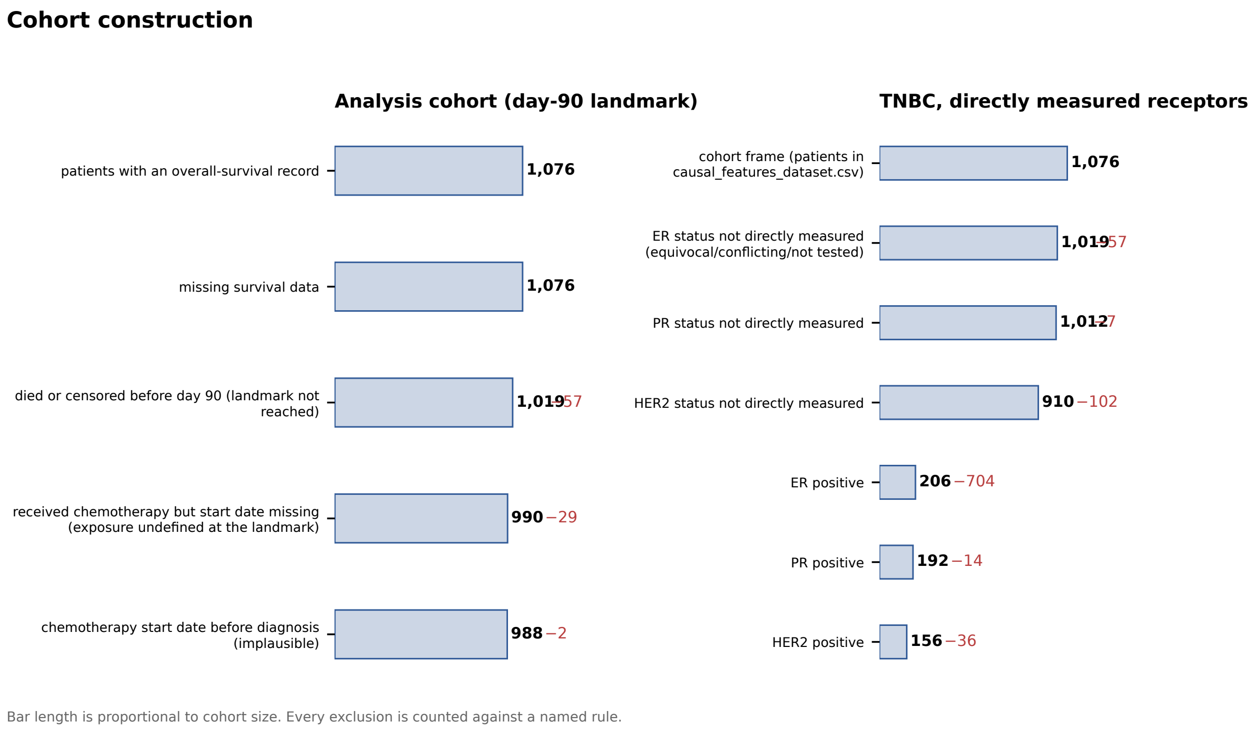


Web Figure 1: Cohort construction and measured TNBC definition. Each exclusion is counted.

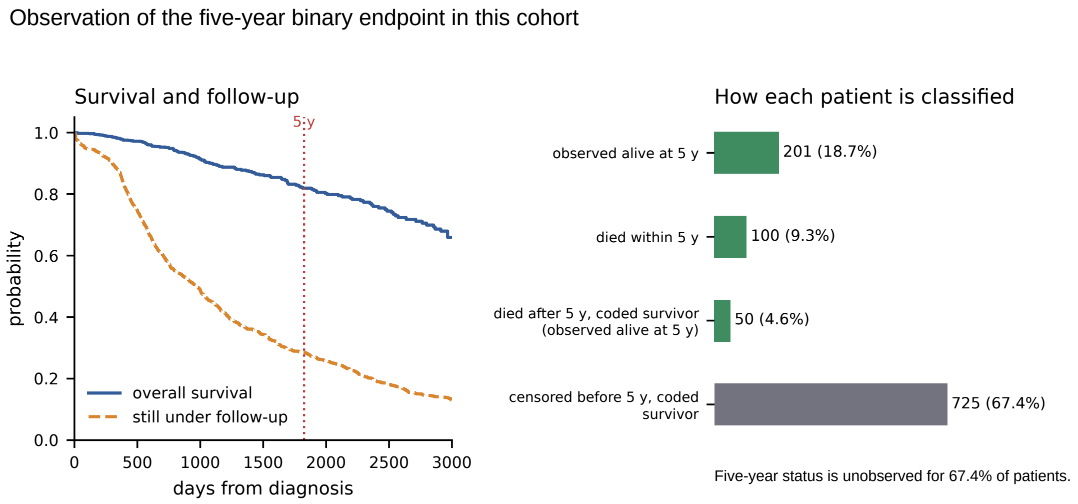


Web Figure 2: Original-cohort survival and follow-up, and ascertainment of the five-year binary endpoint.

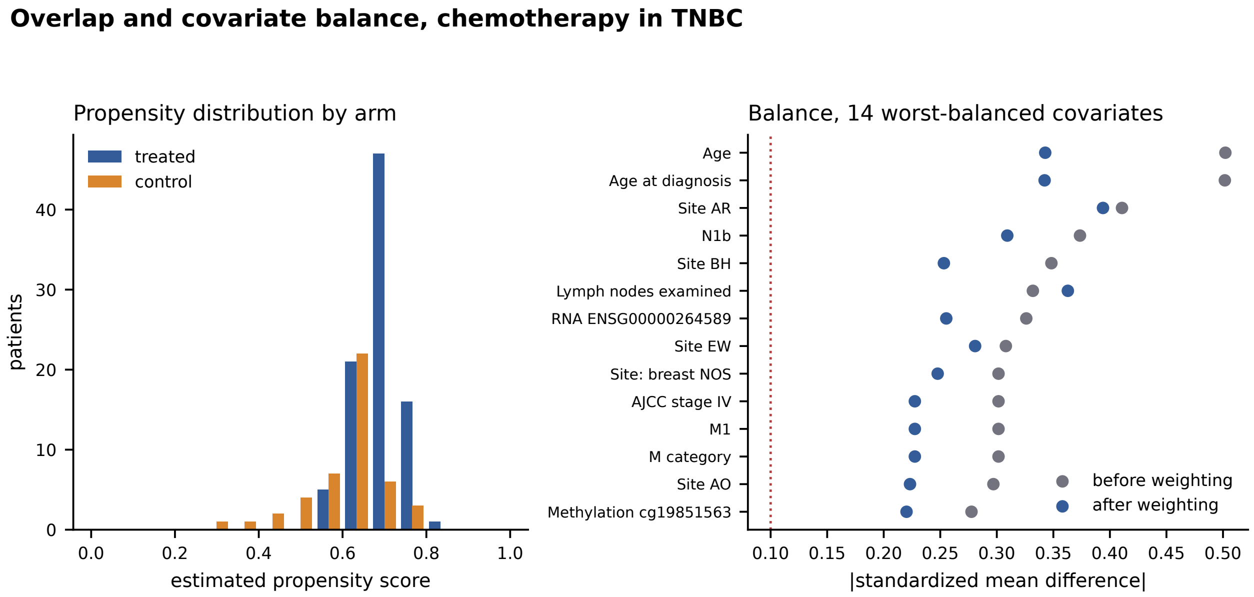


Web Figure 3: Earlier union-set TNBC propensity and balance display, retained as a sensitivity diagnostic. Main-text clinical-primary balance is separately recomputed from the primary estimator weights.

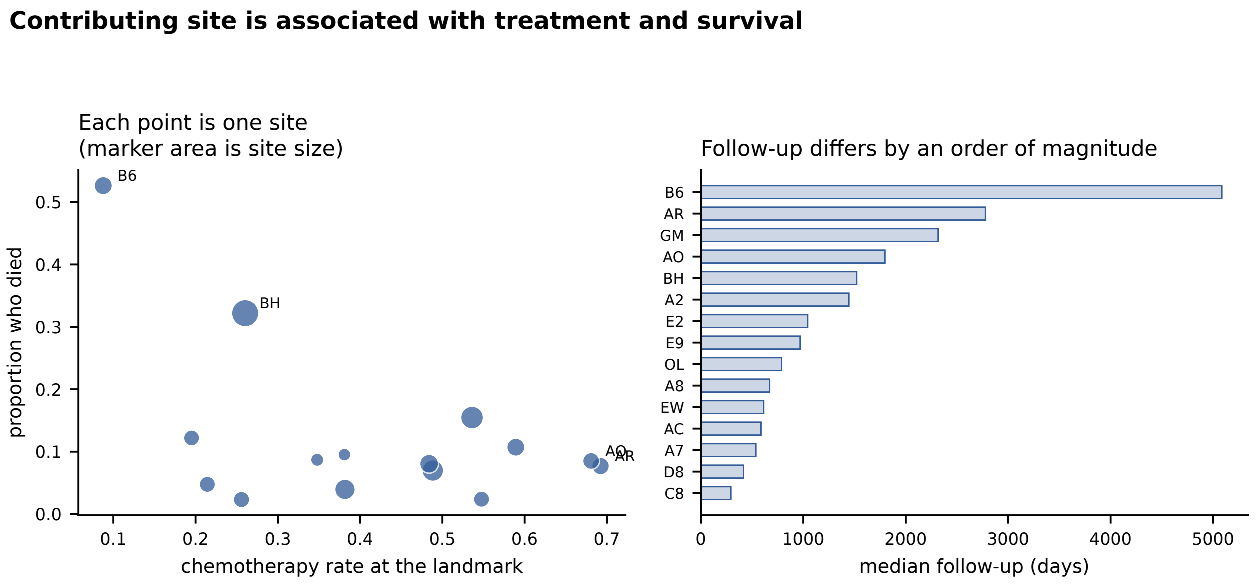


Web Figure 4: Site-level treatment and follow-up summaries. Association alone does not establish that site is a confounder.

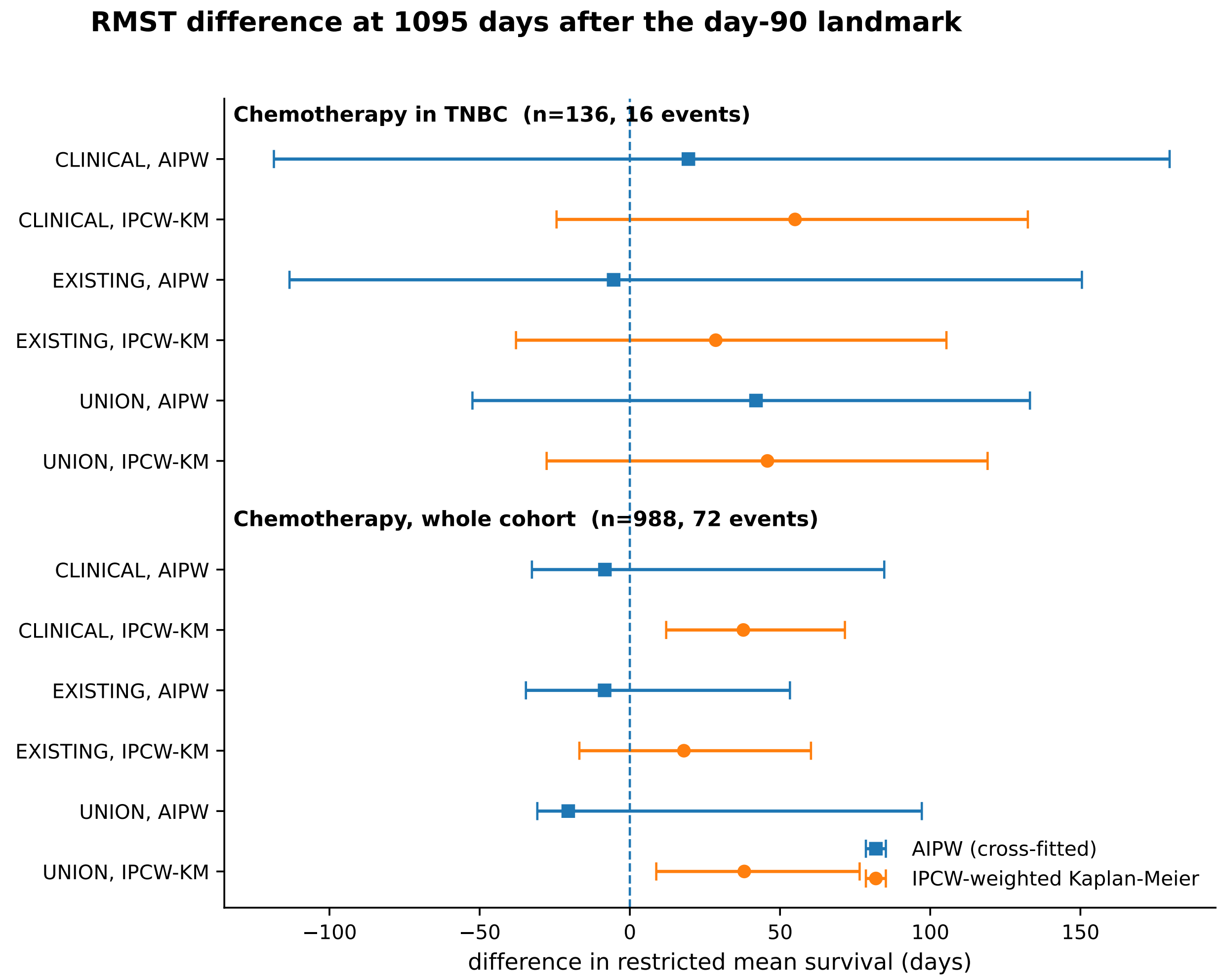


Web Figure 5: Original primary-horizon RMST contrasts under the clinical, existing and union adjustment sets. Bars show 95% patient-bootstrap percentile intervals. This display compares weighted KM and cross-fitted AIPW only; the added unadjusted KM and Cox benchmarks are reported in main Table 2.
